# Frailty in individuals with depression, bipolar disorder and anxiety disorders: longitudinal analyses of all-cause mortality

**DOI:** 10.1101/2022.02.23.22271065

**Authors:** Julian Mutz, Umamah Choudhury, Jinlong Zhao, Alexandru Dregan

**Affiliations:** Social, Genetic and Developmental Psychiatry Centre, Institute of Psychiatry, Psychology & Neuroscience, King’s College London, London, United Kingdom; Department of Basic & Clinical Neuroscience, Institute of Psychiatry, Psychology & Neuroscience, King’s College London, London, United Kingdom; Department of Psychological Medicine, Institute of Psychiatry, Psychology & Neuroscience, King’s College London, London, United Kingdom

**Keywords:** Anxiety disorder, Bipolar disorder, Depression, Frailty, Mortality, UK Biobank

## Abstract

**Background:** Frailty is a medical syndrome that is strongly associated with mortality risk, and an emerging global health burden. Mental disorders are associated with reduced life expectancy and elevated levels of frailty. In this study, we examined the mortality risk associated with frailty in individuals with a lifetime history of mental disorders compared to non-psychiatric controls.

**Methods:** The UK Biobank study recruited >500,000 adults, aged 37–73, between 2006–2010. We derived the two most common albeit distinctive measures of frailty, the frailty phenotype and the frailty index. Individuals with lifetime depression, bipolar disorder or anxiety disorders were identified from multiple data sources. The primary outcome was all-cause mortality. We have also examined differences in frailty, separately by sex and age.

**Outcomes:** Analyses included up to 297,380 middle-aged and older adults with a median follow-up of 12.19 (IQR = 1.31) years, yielding 3,516,706 person-years of follow-up. We observed higher levels of frailty in individuals with mental disorders for both frailty measures. For key comparisons, individuals with a mental disorder had greater all-cause mortality hazards than their controls. The highest hazard ratio (3.65, 95% CI 2.40-5.54) was observed among individuals with bipolar disorder and frailty, relative to the non-frail controls.

**Interpretation:** Our findings highlight elevated levels of frailty across three common mental disorders. The increased mortality risk associated with frailty and mental disorders represents a potentially modifiable target for prevention and treatment to improve life expectancy.

**Funding:** Biotechnology and Biological Sciences Research Council.

## Background

Frailty is a medical syndrome that is characterised by age-related declines in functioning across multiple physiological systems. Frail individuals have a decreased physiological reserve capacity, leaving them less resilient to stressors and at an increased risk of adverse health outcomes. Multiple definitions of frailty exist, most prominently Fried’s frailty phenotype^1^ and the frailty index by Mitnitski, Mogilner and Rockwood^2^. The frailty phenotype comprises five specific indicators of physical capability, while the frailty index represents the accumulation of multiple health deficits across diverse physiological systems and can be adapted to routinely collected data. Frailty is strongly associated with mortality risk^3,4^ and represents an emerging global health burden^5^.

Frailty is also increasingly seen as a valuable clinical measure in psychiatric populations^6^. Individuals with mental disorders are at an increased risk of medical comorbidities^7^, have a lower life expectancy^8^, differ from healthy controls in physiological function^9-11^, and may experience accelerated biological ageing^12^. Frailty is associated with molecular indicators of ageing such as DNA methylation^13^ and provides complementary information to other biomarkers^14,15^. As such, it may be useful for risk stratification^16^ and for predicting adverse health-related outcomes, including disability, falls, loss of independence, and delayed recovery from illness. Frailty represents both a potential mechanism and synergistic factor contributing to the increased mortality risk of individuals with mental disorders.

However, few studies have investigated the mortality risk associated with frailty in adults with mental disorders^17^. As such, the primary aim of this study was to examine the mortality risk associated with frailty in individuals with a lifetime history of mental disorders. Using data from up to 297,380 participants in the UK Biobank, a major biomedical database, we examined all-cause mortality in individuals with depression, bipolar disorder and anxiety disorders. Frailty was assessed using two measures, the Fried frailty phenotype and the frailty index, to enable distinctive yet complementary insights into the impact of frailty on mortality risk in mental disorders. Secondary aims of this study included examining differences in frailty between individuals with mental disorders and non-psychiatric controls, sex-specific effects and age-related changes in frailty.

## Methods

### Study population

The UK Biobank is a prospective study of more than 500,000 middle-aged and older adults who were recruited between 2006 and 2010. The study rationale and design have been described elsewhere^18^. Briefly, individuals registered with the UK National Health Service (NHS) and living within a 25-mile (∼40 km) radius of one of 22 assessment centres were invited to participate. Participants provided data on their sociodemographic characteristics, health behaviours and medical history and underwent physical examinations. Linked hospital inpatient records are available for most participants and primary care records are available for half of the participants. A third of the participants also completed an online follow-up mental health questionnaire (MHQ) between 2016 and 2017.

### Mental disorders

We identified individuals with a lifetime history of depression, bipolar disorder or anxiety disorders using criteria that we have reported elsewhere^9-11^. Cases were ascertained from multiple data sources: the modified Composite International Diagnostic Interview Short Form (CIDI-SF), self-report questions on (hypo)mania and a question on psychiatric diagnoses (UK Biobank data field 20544) which were assessed as part of the MHQ; the nurse-led baseline interview in which participants reported medical diagnoses (field 20002); hospital inpatient records (ICD-10 codes); primary care records (Read v2 or CTV3 codes); self-report questions on mood disorders from the baseline assessment (field 20126). Individuals with psychosis were excluded from all cases and individuals with bipolar disorder were excluded from anxiety disorder cases due to their increased risk of physical multimorbidity^19,20^. The depression and bipolar disorder groups were mutually exclusive, but individuals could be included in both the anxiety disorder and the depression group.

Non-psychiatric controls were defined as individuals who had no mental disorders: (i) had not reported “schizophrenia”, “depression”, “mania / bipolar disorder / manic depression”, “anxiety / panic attacks”, “obsessive compulsive disorder”, “anorexia / bulimia / other eating disorder”, “post-traumatic stress disorder” at the nurse-led interview; (ii) reported no psychiatric diagnoses on the MHQ; (iii) reported no current psychotropic medication use at baseline (field 20003)^21^; (iv) had no ICD-10 Chapter V code in their hospital inpatient record, except for organic causes or substance use; (v) had no diagnostic codes for mental disorders in their primary care record^22^; (vi) were not classified as individuals with probable mood disorder at the baseline assessment; (vii) had no Patient Health Questionnaire-9 (PHQ-9) or Generalised Anxiety Disorder Assessment (GAD-7) sum score of ≥5; (viii) did not report that they ever felt worried, tense, or anxious for most of a month or longer (field 20421); (ix) were not identified as cases based on the CIDI-SF and questions on (hypo)mania^9,11^.

### Frailty phenotype

We derived the Fried frailty phenotype^23^, adapted for the UK Biobank^24,25^. Participants provided data on weight loss, exhaustion, physical activity and walking speed via touch-screen questionnaires at the baseline assessment (Supplement Table 1). Hand-grip strength in whole kilogram-force units was measured using a Jamar J00105 hydraulic hand dynamometer. We used the maximal grip strength of the participant’s self-reported dominant hand. If no data on handedness were available or the participant was ambidextrous, we used the highest value of both hands^26^. All variables were coded as zero or one and summed up. Participants with a total score of three or more were classified as frail, while participants with a total score of one or two and zero were classified as pre-frail and non-frail, respectively^23^. Participants with missing data for at least one criterion were excluded.

### Frailty index

We also derived a frailty index, following the procedure previously used in the UK Biobank^27^. Health deficits included in this index met the following criteria: indicators of poor health; more prevalent in older individuals; neither rare nor universal; covering multiple areas of human functioning; available for ≥80% of participants. The index included 49 variables obtained via touch-screen questionnaires and nurse-led interviews at the baseline assessment, including cardiometabolic, cranial, immunological, musculoskeletal, respiratory and sensory traits, well-being, infirmity, cancer and pain (Supplement Table 2). Categorical variables were dichotomised (no deficit = 0; deficit = 1) and ordinal variables were mapped onto a score between zero and one. The sum of deficits present was divided by the total number of possible deficits, resulting in frailty index values between zero and one, with higher values reflecting greater levels of frailty^28,29^. Participants with missing data for ≥10 variables were excluded^27^. Participants with a frailty index value of ≤0.08 were classified as non-frail, while participants with values between 0.08–0.25 and ≥0.25 were classified as pre-frail and frail, respectively^30^.

### Ascertainment of mortality

The date of death was obtained through linkage with national death registries, NHS Digital (England and Wales) and the NHS Central Register (Scotland). The censoring date was 28 February 2021. The most recent death was recorded for 23 March 2021, although the data were incomplete for March 2021.

### Covariates

Covariates were identified from previous studies and included age, sex, ethnicity (White, Asian, Black, Chinese, Mixed-race or other), highest educational/professional qualification (four levels, reflecting similar years of education^31^: College/University Degree; Education to age 18 or above, but not reaching degree level (“A levels”/”AS levels” or equivalent, NVQ/HND/HNC or equivalent, other professional qualifications); Education to age 16 qualifications (“GCSEs”/”O levels” or equivalent, “CSEs” or equivalent); No qualifications), Townsend deprivation index, which is a small-area level measure of socioeconomic status^32^, cohabitation with spouse or partner (yes/no)^33^, smoking status (never, former or current), alcohol intake frequency (never, special occasions only, one to three times a month, once or twice a week, three or four times a week, or daily or almost daily), systolic and diastolic blood pressure (mmHg), body mass index (BMI, kg/m^2^), cholesterol (mmol/L), multimorbidity count (zero, one, two, three, four, five or more) and assessment centre.

### Statistical analyses

All statistical analyses and data visualisations were done in R (version 3.6.2).

Sample characteristics were summarised using means and standard deviations or counts and percentages. Case-control differences in the frailty index were estimated using standardised mean differences ± 95% confidence intervals (CI) and ordinary least squares regression models. Case-control differences in the frailty phenotype (non-frail, pre-frail and frail) were estimated using ordinal logistic regression models. We fitted both unadjusted and fully adjusted models. Age-related changes in the frailty index were estimated using generalised additive models.

We calculated person-years of follow-up and the median duration of follow-up of censored individuals. Unadjusted survival probabilities by frailty level and case status were estimated using the Kaplan-Meier (KM) method^34^. Hazard ratios (HRs) and 95% confidence intervals were estimated using Cox proportional hazards models^35^ to examine associations between frailty and mortality by case status. Age in years was used as the underlying time axis, with age 40 as the start of follow-up. We fitted both unadjusted and fully adjusted models. Non-frail controls were the reference group. The percentage risk difference between individuals with mental disorders and controls was estimated at the pre-frailty and frailty levels using the formula: (HR_cases_ – HR_controls_)/(HR_controls_ − 1) × 100.

Adjusted *P*-values were calculated using the *p*.*adjust* function in R to account for multiple testing. *P*-values from the regression models were corrected for six tests (one parameter × two models × three disorders) and *p*-values from the Cox proportional hazards models for 30 tests (five parameters × two models × three disorders). Two methods were used: (1) Bonferroni and (2) Benjamini & Hochberg^36^, two-tailed, with *α* = .05 and a 5% false discovery rate, respectively. We have opted for this approach because the Bonferroni correction may be too conservative and lead to a high number of false negatives.

### Additional analyses

We repeated our main analyses of case-control differences in frailty and of all-cause mortality stratified by sex. As a sensitivity analysis, we also repeated the analyses of all-cause mortality excluding individuals with comorbid depression and anxiety disorders.

## Results

The analytical samples included up to 297,380 participants. 76,586 individuals had a lifetime history of depression, 3029 individuals had bipolar disorder and 37,779 individuals had lifetime anxiety disorders. The non-psychiatric control group included 220,794 participants. The percentage of individuals with frailty ranged from 1.8% in the control group to 5.5% in individuals with bipolar disorder. The sample characteristics are presented in Table 1. There was moderate overlap between the frailty phenotype and the frailty index categories (Supplement Figure 1).

**Table 1.** Sample characteristics of individuals with mental disorders and non-psychiatric controls

|  | Depression<br>N=76586 | Bipolar disorder<br>N=3029 | Anxiety disorder<br>N=37779 | Controls<br>N=220794 |
| --- | --- | --- | --- | --- |
| **Age** |  |  |  |  |
| Mean (SD) | 55.13 (7.92) | 54.31 (8.00) | 55.72 (7.89) | 56.43 (8.15) |
| **Sex** |  |  |  |  |
| Female | 50040 (65.3%) | 1710 (56.5%) | 24944 (66.0%) | 109447 (49.6%) |
| Male | 26546 (34.7%) | 1319 (43.5%) | 12835 (34.0%) | 111347 (50.4%) |
| **Neighbourhood deprivation** |  |  |  |  |
| Mean (SD) | -1.48 (2.91) | -1.10 (3.02) | -1.62 (2.87) | -1.78 (2.83) |
| **Ethnicity** |  |  |  |  |
| White | 73868 (96.5%) | 2855 (94.3%) | 36754 (97.3%) | 209251 (94.8%) |
| Mixed-race | 522 (0.7%) | 28 (0.9%) | 191 (0.5%) | 1105 (0.5%) |
| Black | 613 (0.8%) | 41 (1.4%) | 248 (0.7%) | 3280 (1.5%) |
| Asian | 955 (1.2%) | 67 (2.2%) | 351 (0.9%) | 4521 (2.0%) |
| Chinese | 120 (0.2%) | 6 (0.2%) | 42 (0.1%) | 822 (0.4%) |
| Other | 508 (0.7%) | 32 (1.1%) | 193 (0.5%) | 1815 (0.8%) |
| **Highest qualification** |  |  |  |  |
| None | 10642 (13.9%) | 316 (10.4%) | 5197 (13.8%) | 35669 (16.2%) |
| O levels/GCSEs/CSEs | 21309 (27.8%) | 794 (26.2%) | 10465 (27.7%) | 61512 (27.9%) |
| A levels/NVQ/HND/HNC<sup>1</sup> | 18105 (23.6%) | 714 (23.6%) | 8931 (23.6%) | 51268 (23.2%) |
| Degree | 26530 (34.6%) | 1205 (39.8%) | 13186 (34.9%) | 72345 (32.8%) |
| **Spouse/partner cohabitation** |  |  |  |  |
| No | 10864 (14.2%) | 521 (17.2%) | 4736 (12.5%) | 19148 (8.7%) |
| Yes | 65722 (85.8%) | 2508 (82.8%) | 33043 (87.5%) | 201646 (91.3%) |
| **Smoking status** |  |  |  |  |
| Never | 39622 (51.7%) | 1408 (46.5%) | 19868 (52.6%) | 126857 (57.5%) |
| Former | 28321 (37.0%) | 1122 (37.0%) | 14128 (37.4%) | 75365 (34.1%) |
| Current | 8643 (11.3%) | 499 (16.5%) | 3783 (10.0%) | 18572 (8.4%) |
| **Alcohol intake frequency** |  |  |  |  |
| Never | 6242 (8.2%) | 329 (10.9%) | 3163 (8.4%) | 14311 (6.5%) |
| Special occasions | 9487 (12.4%) | 410 (13.5%) | 4552 (12.0%) | 21354 (9.7%) |
| 1-3/month | 9314 (12.2%) | 352 (11.6%) | 4256 (11.3%) | 22680 (10.3%) |
| 1-2/week | 19101 (24.9%) | 724 (23.9%) | 9271 (24.5%) | 59197 (26.8%) |
| 3-4/week | 16966 (22.2%) | 578 (19.1%) | 8554 (22.6%) | 55923 (25.3%) |
| Daily/almost daily | 15476 (20.2%) | 636 (21.0%) | 7983 (21.1%) | 47329 (21.4%) |
| **Body mass index** |  |  |  |  |
| Mean (SD) | 27.68 (5.07) | 28.15 (5.37) | 27.36 (4.96) | 27.19 (4.44) |
| **Systolic blood pressure** |  |  |  |  |
| Mean (SD) | 135.00 (18.15) | 133.79 (17.61) | 135.89 (18.34) | 138.64 (18.61) |
| **Diastolic blood pressure** |  |  |  |  |
| Mean (SD) | 81.54 (10.06) | 81.53 (10.15) | 81.66 (10.02) | 82.50 (10.06) |
| **Cholesterol** |  |  |  |  |
| Mean (SD) | 5.72 (1.14) | 5.67 (1.15) | 5.74 (1.13) | 5.70 (1.13) |
| **Multimorbidity count** |  |  |  |  |
| None | 12689 (16.6%) | 380 (12.5%) | 5930 (15.7%) | 62516 (28.3%) |
| One | 18249 (23.8%) | 651 (21.5%) | 8790 (23.3%) | 62660 (28.4%) |
| Two | 15760 (20.6%) | 620 (20.5%) | 7939 (21.0%) | 43558 (19.7%) |
| Three | 11656 (15.2%) | 516 (17.0%) | 5934 (15.7%) | 25471 (11.5%) |
| Four | 7485 (9.8%) | 342 (11.3%) | 3784 (10.0%) | 13374 (6.1%) |
| Five or more | 10747 (14.0%) | 520 (17.2%) | 5402 (14.3%) | 13215 (6.0%) |
| **Antidepressant use** |  |  |  |  |
| No | 60326 (78.8%) | 2175 (71.8%) | 29869 (79.1%) | 220794 (100.0%) |
| Yes | 16260 (21.2%) | 854 (28.2%) | 7910 (20.9%) | 0 (0.0%) |
| **Antipsychotic use** |  |  |  |  |
| No | 76276 (99.6%) | 2827 (93.3%) | 37586 (99.5%) | 220794 (100.0%) |
| Yes | 310 (0.4%) | 202 (6.7%) | 193 (0.5%) | 0 (0.0%) |
| **Lithium use** |  |  |  |  |
| No | 76486 (99.9%) | 2755 (91.0%) | 37740 (99.9%) | 220794 (100.0%) |
| Yes | 100 (0.1%) | 274 (9.0%) | 39 (0.1%) | 0 (0.0%) |
| **Frailty phenotype** |  |  |  |  |
| Non-frail | 35473 (46.3%) | 1284 (42.4%) | 18007 (47.7%) | 125980 (57.1%) |
| Pre-frail | 37549 (49.0%) | 1579 (52.1%) | 18199 (48.2%) | 90881 (41.2%) |
| Frail | 3564 (4.7%) | 166 (5.5%) | 1573 (4.2%) | 3933 (1.8%) |
| **Frailty phenotype count** |  |  |  |  |
| None | 35473 (46.3%) | 1284 (42.4%) | 18007 (47.7%) | 125980 (57.1%) |
| One | 27560 (36.0%) | 1143 (37.7%) | 13578 (35.9%) | 72679 (32.9%) |
| Two | 9989 (13.0%) | 436 (14.4%) | 4621 (12.2%) | 18202 (8.2%) |
| Three | 2870 (3.7%) | 134 (4.4%) | 1264 (3.3%) | 3376 (1.5%) |
| Four | 632 (0.8%) | 30 (1.0%) | 277 (0.7%) | 523 (0.2%) |
| Five | 62 (0.1%) | 2 (0.1%) | 32 (0.1%) | 34 (0.0%) |
| Mean (SD) | 0.77 (0.88) | 0.84 (0.91) | 0.74 (0.86) | 0.55 (0.73) |
| **Frailty index** |  |  |  |  |
| Mean (SD) | 0.15 (0.08) | 0.17 (0.08) | 0.15 (0.08) | 0.10 (0.06) |
| **Frailty index categories** |  |  |  |  |
| Non-frail | 14397 (18.8%) | 445 (14.7%) | 6641 (17.6%) | 88189 (39.9%) |
| Pre-frail | 53280 (69.6%) | 2098 (69.3%) | 26581 (70.4%) | 125648 (56.9%) |
| Frail | 8909 (11.6%) | 486 (16.0%) | 4557 (12.1%) | 6957 (3.2%) |

*Note:* SD = standard deviation; GCSEs = general certificate of secondary education; CSE = certificate of secondary education; NVQ = national vocational qualification; HND = higher national diploma; HNC = higher national certificate. <sup>1</sup> also includes 'other professional qualifications'. Cut-offs for frailty index categories were non frail ( $\leq 0.08$ ), pre-frail ( $>0.08$ and $<0.25$ ) and frail ( $\geq 0.25$ ).

The percentage of participants with pre-frailty or frailty was higher in individuals with mental disorders than in the control group (Table 1). We observed a similar pattern for the frailty criteria count, showing that the percentage of individuals with mental disorders was higher than the percentage of controls for scores 1 to 5 (Figure 1). The largest difference was observed between individuals with bipolar disorder and the control group.

**Figure 1.**
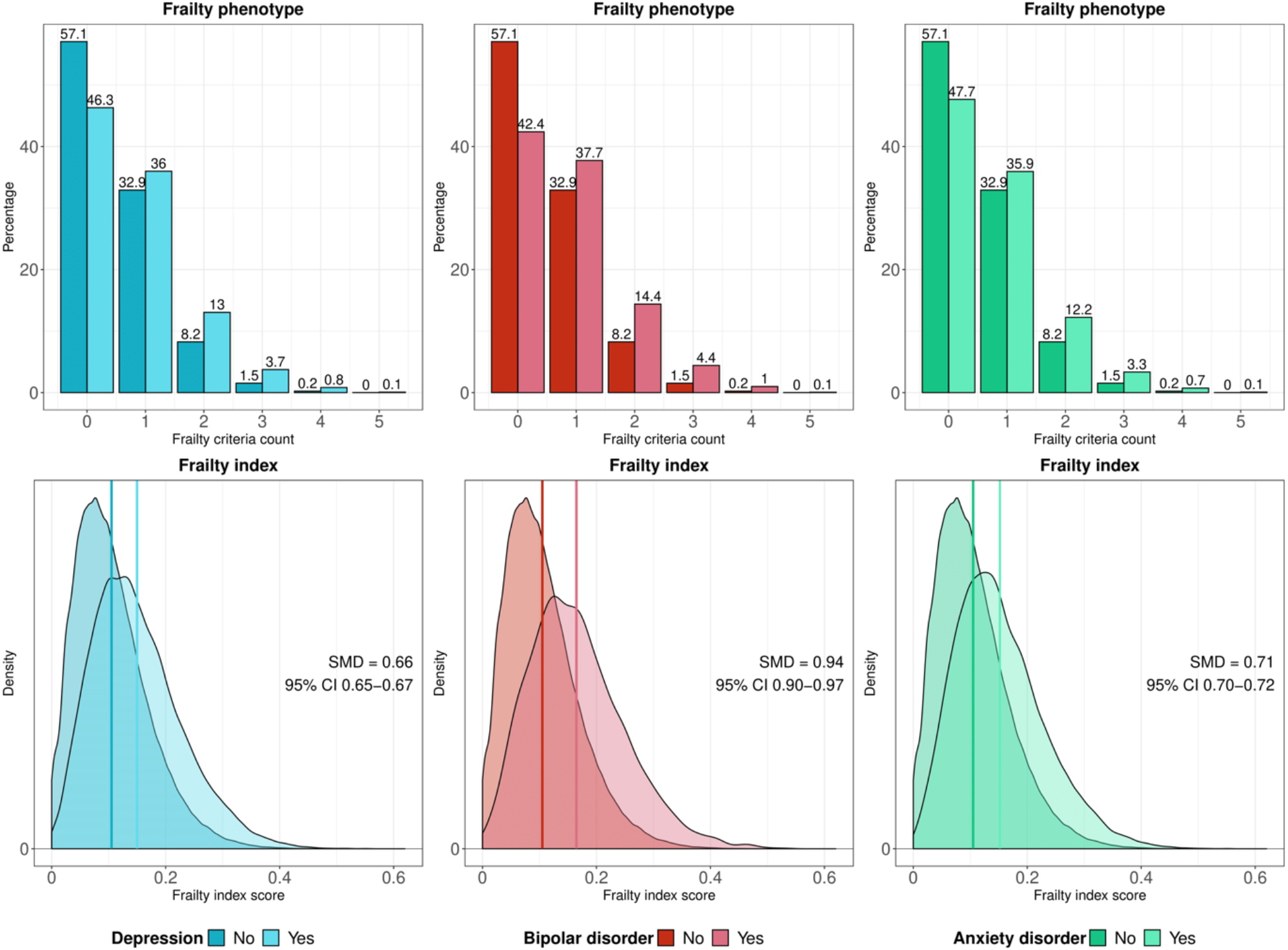
Histograms and density plots showing the distribution of the frailty phenotype criteria count (top panels) and the frailty index (bottom panels), respectively, for individuals with mental disorders and non-psychiatric controls.

Frailty index scores were also higher in individuals with mental disorders, with the largest standardised mean difference (SMD) observed between individuals with bipolar disorder and the controls (SMD = 0.94 (95% CI 0.90-0.97, *p* < 0.001) (Figure 1). Individuals with mental disorders also had higher levels of frailty after adjustment for potential confounders in regression models, irrespective of how frailty was measured (Figure 2 and Table 2).

**Figure 2.**
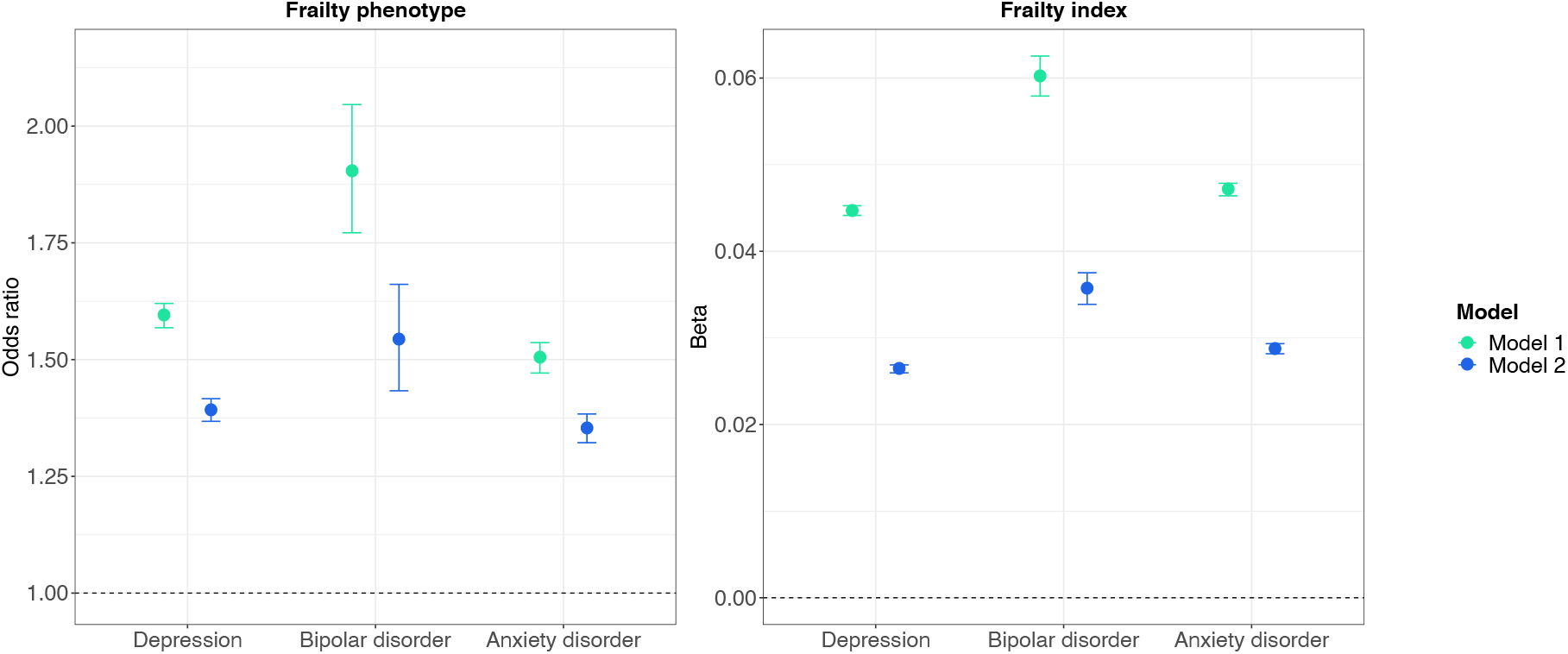
Frailty in individuals with mental disorders compared to non-psychiatric controls (reference group). Estimates shown for the frailty phenotype are odds ratios and 95% confidence intervals (CI) from ordinal logistic regression models, indicating changes in odds of being frailer associated with being in the case group relative to the control group. Estimates shown for the frailty index are ordinary least squares regression beta coefficients and 95% CI. Model 1 – unadjusted; Model 2 – adjusted for age, sex, ethnicity, highest qualification, Townsend deprivation index, cohabitation with spouse/partner, smoking status, alcohol intake frequency, systolic and diastolic blood pressure, body mass index, cholesterol, multimorbidity count and assessment centre.

**Table 2.** Frailty in individuals with mental disorders and non-psychiatric controls

|  | Model 1 |  |  | Model 2 |  |  |
| --- | --- | --- | --- | --- | --- | --- |
| **Frailty phenotype** | OR | 95% CI |  | OR | 95% CI |  |
| Controls | Ref | - | - | Ref | - | - |
| Depression | 1.594 | 1.568 | 1.620 | 1.392 | 1.368 | 1.416 |
| Bipolar disorder | 1.904 | 1.772 | 2.046 | 1.543 | 1.433 | 1.661 |
| Anxiety disorder | 1.503 | 1.471 | 1.536 | 1.352 | 1.322 | 1.384 |
| **Frailty index** | $\beta$ | 95% CI | | $\beta$ | 95% CI | |
| Controls | Ref | - | - | Ref | - | - |
| Depression | 0.045 | 0.044 | 0.045 | 0.026 | 0.026 | 0.027 |
| Bipolar disorder | 0.060 | 0.058 | 0.063 | 0.036 | 0.034 | 0.038 |
| Anxiety disorder | 0.047 | 0.046 | 0.048 | 0.029 | 0.028 | 0.029 |

*Note:* OR = odds ratio; $\beta$ = ordinary least squares regression beta coefficient; CI = confidence interval; Ref = reference group. All Bonferroni-adjusted $p$ -values < 0.001. Odds ratios indicate changes in odds of being frailer associated with being in the case group relative to the control group. Model 1 – unadjusted; Model 2 – adjusted for age, sex, ethnicity, highest qualification, Townsend deprivation index, cohabitation with spouse/partner, smoking status, alcohol intake frequency, systolic and diastolic blood pressure, body mass index, cholesterol, multimorbidity count and assessment centre.

Sample characteristics stratified by sex are presented in Supplement Table 3. Females with mental disorders had higher levels of pre-frailty and frailty than males. A similar pattern emerged with respect to the frailty criteria count, although few individuals had a score of 5 (Supplement Figure 2). Frailty index scores were greater among females compared to males with anxiety disorders, but the magnitude of this difference was negligible compared to the differences between individuals with mental disorders and non-psychiatric controls. There was no evidence of a difference in frailty index scores between males and females with depression or bipolar disorder (Supplement Table 4). In the sex-stratified regression models, both males and females with mental disorders had higher levels of frailty than the controls, including after adjustment for potential confounders (Supplement Figure 3 and Supplement Table 5). For the frailty phenotype, estimates for females were slightly higher than for males, relative to the control group, while we observed mostly the reverse pattern for the frailty index. However, the magnitude of these differences in estimates was negligible.

Frailty index scores increased with age in individuals with mental disorders and in the non-psychiatric control group. We found some evidence that the group differences in frailty between individuals with and without mental disorders narrowed above age 60, resulting from a steeper age-related increase in frailty in the control group (Figure 3).

**Figure 3.**
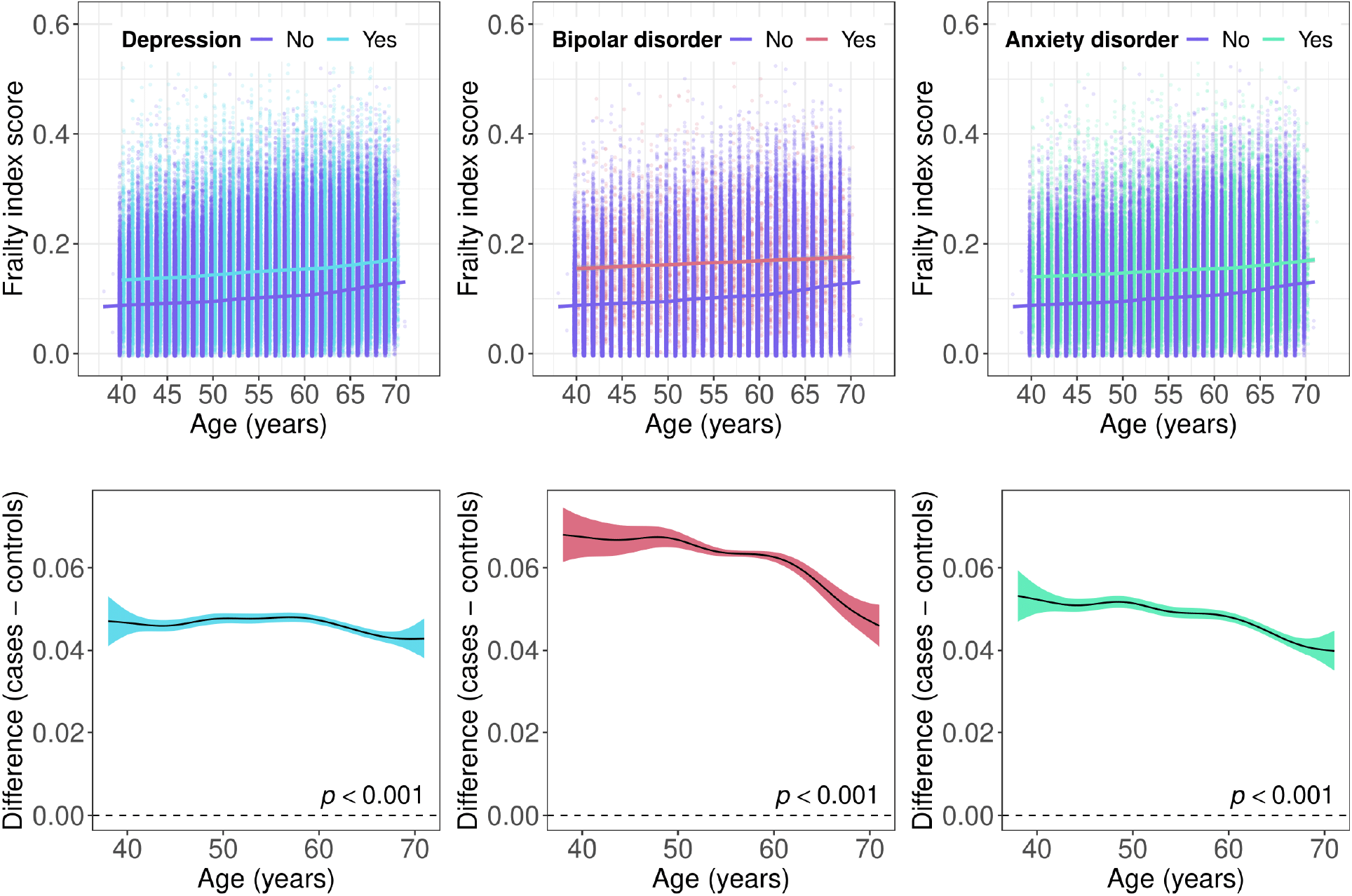
Top panels: scatter plots showing the frailty index by age in individuals with mental disorders and non-psychiatric controls. Bottom panels: difference smooths comparing age-related changes in frailty index scores of individuals with mental disorders to non-psychiatric controls. Positive values on the y-axes correspond to higher frailty index scores in individuals with mental disorders. The smooth curves were estimated using generalised additive models. The shaded areas correspond to 95% confidence intervals.

Differences in the frailty phenotype between individuals with and without mental disorders were fairly consistent across the age spectrum. The combined percentage of participants with pre-frailty or frailty was greater in individuals with mental disorders at most ages, with median estimates of 53.72% in depression, 57.14% in bipolar disorder, 53.65% in anxiety disorders and 43.37% in the control group (Supplement Figure 4).

### All-cause mortality

The median duration of follow-up of censored individuals was between 12.09 (IQR = 1.35) and 12.19 (IQR = 1.31) years, with 2,654,566 to 3,516,706 person-years of follow-up (Supplement Table 6). Individuals with depression or bipolar disorder had a greater all-cause mortality hazard than non-psychiatric controls, while we did not observe an increased mortality risk in individuals with anxiety disorders (Supplement Figure 5). Regardless of frailty measure, the hazards for all-cause mortality were greater among pre-frail and frail participants (Supplement Figure 6). Survival probabilities by frailty level and case status are presented in Supplement Figures 7 and 8.

Considering the frailty phenotype, the largest hazard ratio (HR) was observed for individuals with bipolar disorder and frailty (HR = 3.65, 95% CI 2.40-5.54) compared to non-frail controls (Table 3). A similar pattern of results was revealed for depression and anxiety disorders, although the differences in all-cause mortality were not statistically significant between non-frail individuals with depression or anxiety disorders and non-frail controls. Adjustment for potential confounders attenuated the effect sizes, but the differences persisted (Supplement Figure 9). Results from the Cox proportional hazards models in which we examined the frailty index categories suggested further differences from the analysis of the frailty phenotype (Table 4). For instance, several estimates suggested lower all-cause mortality hazards in individuals with depression or anxiety disorders, relative to their controls, both in the pre-frail and frail groups (Supplement Figure 10).

**Table 3.**
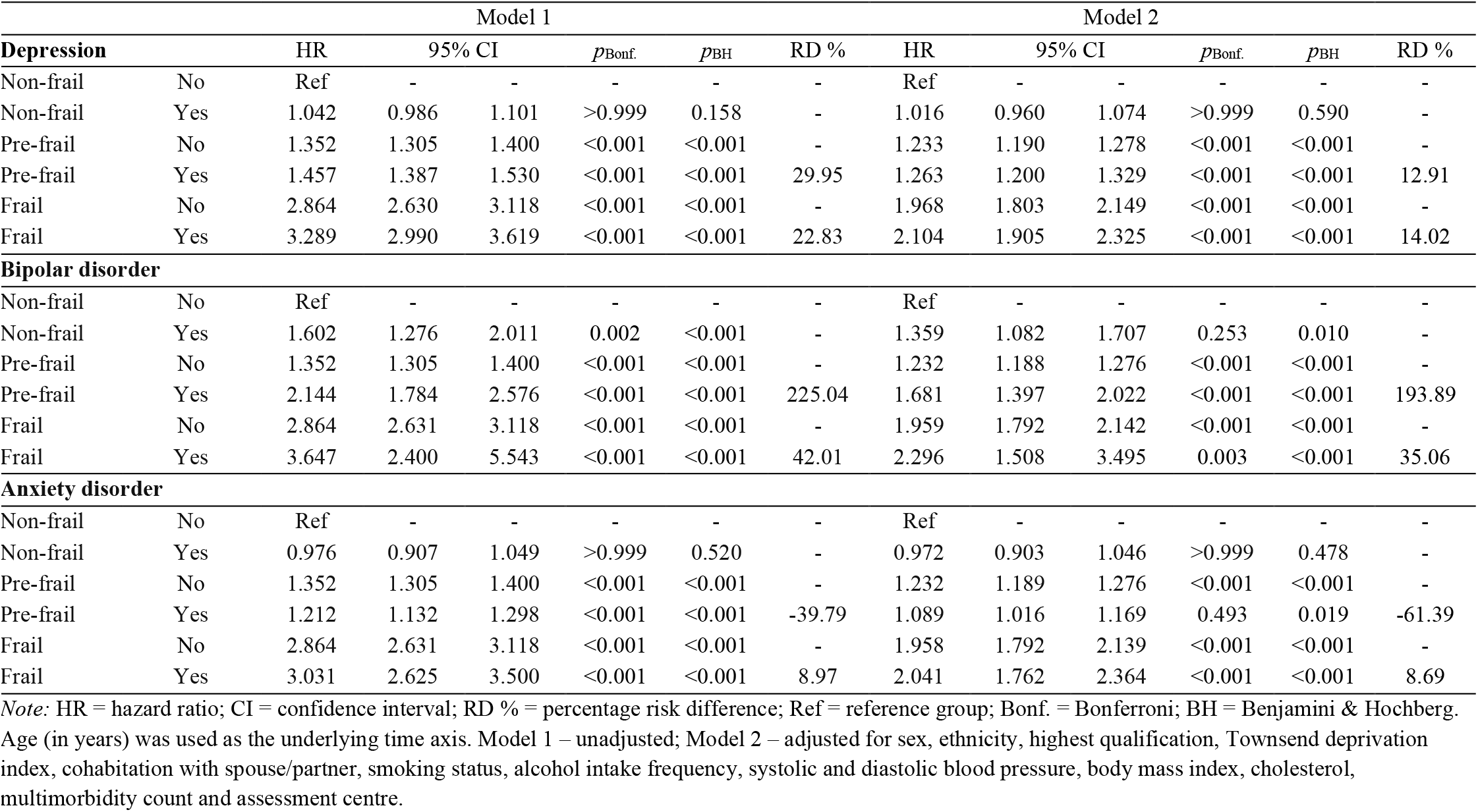
All-cause mortality by frailty phenotype in individuals with mental disorders and non-psychiatric controls

**Table 4.**
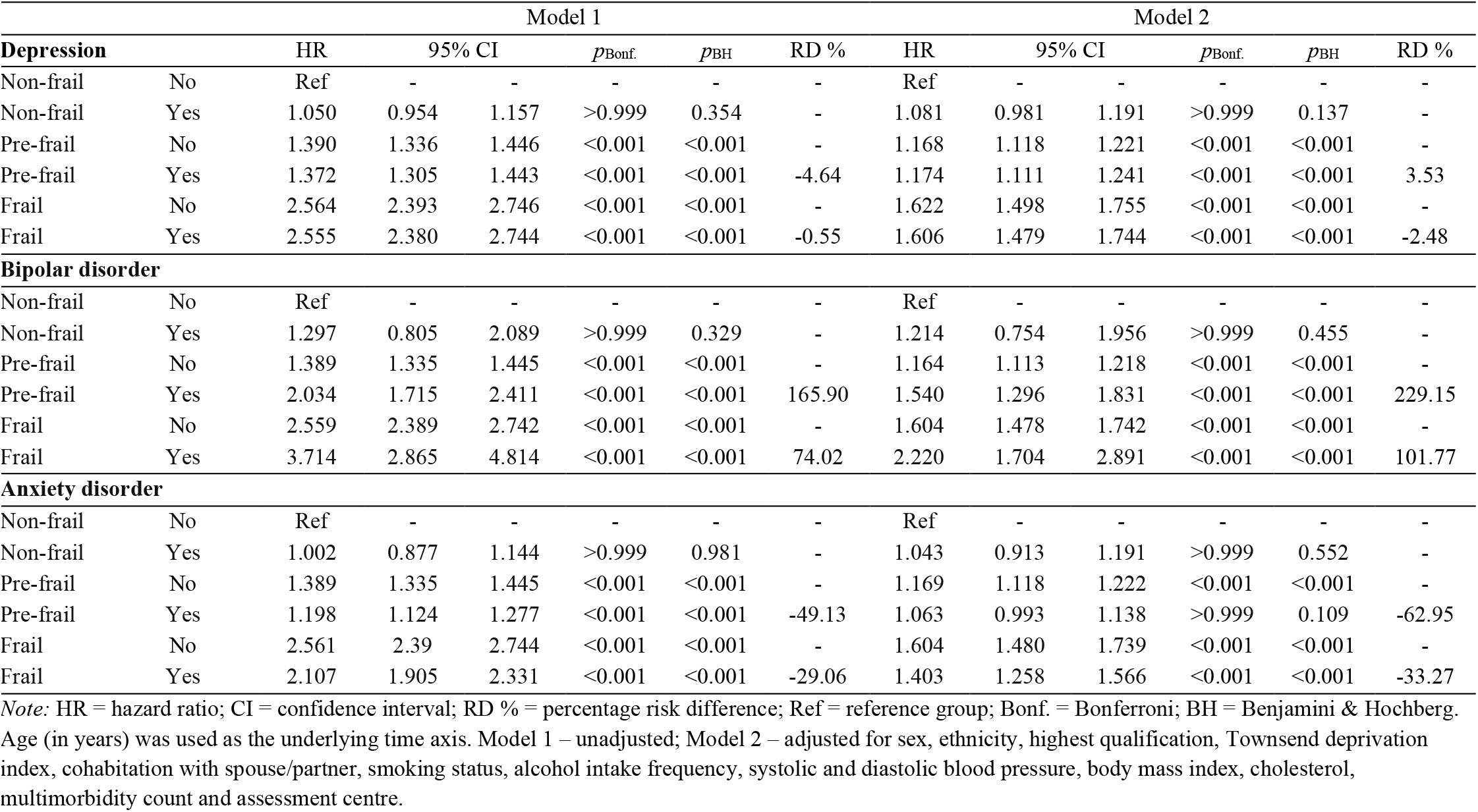
All-cause mortality by frailty index categories in individuals with mental disorders and non-psychiatric controls

### Additional analyses

The results of the analyses of the frailty phenotype and all-cause mortality stratified by sex are presented in Supplement Figure 11 and Supplement Table 7. Overall, males presented with modestly greater all-cause mortality hazards relative to females. Compared to the non-frail control group, pre-frail females with depression had a higher risk of all-cause mortality (though the effect size was lower relative to the pre-frail females in the control group). Frail males with bipolar disorder had a greater all-cause mortality hazard compared to females (adjusted HR = 3.11, 95% CI 1.87-5.18 and HR = 1.39, 95% CI 0.66-2.92, respectively), relative to non-frail controls, while the reverse was observed for the pre-frail groups. Moreover, frail females with anxiety disorders presented with higher all-cause mortality hazards than their male counterparts, relative to the non-frail controls. Of note, frail males with anxiety disorders had a somewhat lower risk of all-cause mortality relative to their counterparts in the control group (adjusted HR = 1.87, 95% CI 1.48-2.35 and HR = 2.01, 95% CI 1.79-2.26, respectively). The overall pattern of results from the sex-stratified models of the frailty index categories mirrored the results from the main analysis (Supplement Figure 12 and Supplement Table 8). Pre-frail females with bipolar disorder, anxiety disorders or in the control group had a higher mortality hazard than males, relative to the non-frail control group. The mortality hazard of non-frail females with bipolar disorder was also higher than in males.

The results for the subset of participants from which we excluded individuals with comorbid depression and anxiety disorders (*n* = 23,712) are shown in Supplement Tables 9-11. Overall, the all-cause mortality hazards were slightly elevated in these analyses.

## Discussion

In a large prospective study, we observed higher levels of frailty in individuals with mental disorders compared to people without mental disorders, regardless of how frailty was defined. The frailty phenotype measure was consistent in documenting higher pre-frailty and frailty levels in females compared to males with mental disorders. Evidence for sex differences in the frailty index was observed mainly within anxiety disorders, with females demonstrating higher frailty scores relative to males. Notably, our findings suggested that differences in the frailty index scores between individuals with mental disorders and non-psychiatric controls narrowed above age 60. On the other hand, there were mostly consistent differences in the frailty phenotype (both at the pre-frail and frail levels) between individuals with mental disorders and their controls across the age spectrum.

The above differences in frailty levels translated into an increased risk of all-cause mortality among individuals with a lifetime history of depression or bipolar disorders with respect to both the frailty phenotype and the frailty index measures. The association between the frailty index and anxiety disorders with all-cause mortality was less consistent. Concerning sex differences, our findings revealed increased all-cause mortality among males relative to females, with certain exceptions. For instance, pre-frail males with bipolar disorder and frail males with anxiety disorders appeared to have a lower risk of all-cause mortality relative to females.

Previous studies that have examined differences in frailty between individuals with and without mental disorders have focussed on depression and older adults^37,38^. A meta-analysis of 24 cross-sectional and longitudinal studies suggested increased frailty levels among people with depression^38^. Our study findings supported this evidence and extended it to individuals with bipolar disorder or anxiety disorders. Our finding that differences in the frailty index between individuals with mental disorders and controls narrowed with age is consistent with a previous study showing that the relationship between depressive symptoms and the frailty phenotype weakened as people aged^39^. A potential explanation for this could be better coping strategies in older individuals. The lack of evidence of age-related increases in the frailty phenotype may be due to the younger age of the participants in our study, as previous research in adults aged 65 years or older observed increased frailty at older ages^40^. It could also be due to selection bias resulting in healthier older adults participating at greater rates. Recently we have observed a decline in the prevalence of common mental disorders among adults in their late 50s to early 60s, followed by a sharp increase afterwards.^41^ This trend represents another possible explanation for the decline in the strength of the association between the frailty phenotype and mental disorders in individuals over 60 years of age observed in the current study.

The dose-response association between the frailty phenotype levels and mortality that we observed in this study is consistent with a meta-analysis of population-based studies that included >35,000 adults aged 65 years and above^3^. However, there has been little research to date examining the mortality risk associated with frailty in individuals with specific mental disorders. Findings from a prospective study of 2565 men aged 75 years or older suggested that current symptoms of depression, but not lifetime depression, were associated with increased all-cause mortality, and that this association was largely due to differences in frailty^42^. Another recent small study (*N* = 378) of patients with depression aged 60 years or older documented that the frailty phenotype count was associated with increased mortality risk^17^. Another small study (*N* = 120) with older adults who were admitted for psychiatric inpatient treatment suggested that frailty was a strong predictor of mortality within this population^43^. However, this study did not provide data on disorder-specific mortality rates associated with frailty. Finally, a previous study of multimorbidity and frailty suggested that individuals with neuropsychiatric multimorbidity had the highest mortality rate for each level of frailty^44^. To the best of our knowledge, our study is the first to examine the mortality risk associated with frailty in individuals with bipolar disorder or anxiety disorders.

The inclusion of distinctive indicators of frailty enabled us to provide more robust evidence about the role of frailty in premature mortality among a large group of people with mental disorders. The identification of subpopulations at risk of accelerated physiological decline is informative for the implementation of preventative strategies aimed at reducing the excess mortality in individuals with mental disorders. In general, physical frailty arises from dysregulation in multiple and dynamic body systems over long periods of time^45^. Individually tailored multicomponent interventions (e.g., physical activity, diet, psycho-social support and integrated care models) are likely to offer the best prognosis for ameliorating frailty within mental health populations. The evidence for the efficacy of such interventions to modify frailty in individuals with mental disorders is currently limited. In the meantime, the focus should be on minimising potentially aggravating factors for frailty in people with mental disorders, such as inappropriate polypharmacy, lack of care continuity, or social isolation, while optimising integrated care and healthy behaviours (e.g., physical activity)^46^. Further longitudinal studies are needed to understand how the progression of frailty impacts on the progression of mental disorders and vice versa. In addition, further studies using multisystem physiological markers of frailty may help detect the inflection point for frailty-related mortality in people with mental disorders.

A strength of this study is the large sample size of almost 300,000 participants with a median follow-up of 12 years. We focussed on two distinctive yet complementary measures of frailty, the frailty phenotype and frailty index, and examined their association with all-cause mortality among three mental disorders with considerable disease burden. While research on frailty has predominantly been conducted in individuals aged 65 years and above, our study sample included both middle-aged and older adults, highlighting the association of frailty with all-cause mortality at the transition from middle age to late adulthood years.

Our observational research has certain limitations. As we have discussed elsewhere^47,48^, UK Biobank participants are healthier than the UK general population. As such, individuals with high levels of frailty and with chronic and/or severe mental disorders may have been less likely to be included in our study. This could have resulted in attenuated case-control differences in frailty and reduced the corresponding mortality risk. For a discussion of the potential limitations regarding the ascertainment of individuals with mental disorders in the UK Biobank, see our previous studies^9-11^. There is a conceptual overlap between frailty and some of the symptoms that characterise mental disorders. However, a previous study of community-dwelling older adults showed that shared symptoms explained only part of the association between depression and frailty^49^. Our study provides limited insight into the causal relationships between mental disorders and frailty in relation to mortality. However, it is likely that mental illness and frailty are mutually reinforcing, and may share common risk factors^50^. Finally, we cannot exclude the possibility of residual confounding, and other variables (e.g., genetics, healthcare access or drug prescriptions) not considered here may also affect the observed associations. Our estimation models did adjust for a wide range of known confounders, however, minimising the potential risk from residual bias.

### Conclusion

Our findings highlight elevated levels of frailty across three common mental disorders. Screening for frailty might help identify individuals with mental disorders who are at risk of premature mortality. Screening for poor mental health is equally important as mental disorders tend to be under-recognised in individuals presenting with high levels of frailty and physical comorbidities. There is increasing evidence that frailty can be prevented, treated and potentially delayed. The increased mortality risk associated with frailty and mental disorders represents a modifiable target to increase healthy life expectancy, especially where frailty and mental disorders coexist.

## Research in context

### Evidence before this study

Frailty is an emerging global health burden and associated with increased mortality risk. We searched the Embase, MEDLINE, Global Health and APA PsycINFO electronic databases for articles published between 2001 and 2022, using the following combination of search terms: (“frailty” OR “frail$”) AND (“depress$” OR “bipolar” OR “bipolar disorder” OR “anxiety” OR “anxiety disorder”) AND (“mortality” OR “death”). We found that individuals with mental disorders had higher levels of frailty. There has been little previous research that examined how frailty predicts all-cause mortality in individuals with lifetime depression, bipolar disorder or anxiety disorders.

### Added value of this study

Our findings highlight elevated levels of frailty across three common mental disorders in a large population-scale study. Our study is the first to examine the mortality risk associated with distinctive yet complementary measures of frailty in individuals with bipolar disorder or anxiety disorders. Middle-aged and older adults with mental disorders and frailty generally had the highest mortality risk, with the greatest mortality hazard observed in individuals with bipolar disorder and frailty.

### Implications of all the available evidence

The increased mortality risk associated with frailty and mental disorders represents a potentially modifiable target for prevention and treatment to improve life expectancy, especially where frailty and mental disorders coexist. While waiting for more definitive trials on multicomponent interventions, the focus of clinical practice should concern minimising the adverse impact of inappropriate polypharmacy, while optimising integrated care and resilience-promoting behaviours, such as physical activity.

## Data Availability

The data used are available to all bona fide researchers for health-related research that is in the public interest, subject to an application process and approval criteria. Study materials are publicly available online at http://www.ukbiobank.ac.uk.

## Ethics

Ethical approval for the UK Biobank study has been granted by the National Information Governance Board for Health and Social Care and the NHS North West Multicentre Research Ethics Committee (11/NW/0382). No project-specific ethical approval is needed. Data access permission has been granted under UK Biobank application 45514. Written informed consent was obtained from all participants.

## Acknowledgements

This research has been conducted using data from UK Biobank, a major biomedical database. This project made use of time on Rosalind HPC, funded by Guy’s & St Thomas’ Hospital NHS Trust Biomedical Research Centre (GSTT-BRC), South London & Maudsley NHS Trust Biomedical Research Centre (SLAM-BRC), and Faculty of Natural Mathematics & Science (NMS) at King’s College London.

## Author Contribution

JM conceived the idea of the study, acquired the data, carried out the statistical analysis, interpreted the findings, wrote the manuscript and revised the manuscript for final submission. AD contributed to the study design, interpreted the findings and revised the manuscript. UC and JZ contributed to the data pre-processing and reviewed the manuscript. All authors read and approved the final manuscript.

## Supplementary material

Supplementary information is available online.

