## Supplement for "Frailty in individuals with depression, bipolar disorder and anxiety disorders: longitudinal analyses of all-cause mortality"

This material accompanies the article

#### Full criteria used to identify individuals with mental disorders

Criteria below are adapted from Mutz and Lewis (2021)[1] Mutz et al. (2022)[2] and Mutz et al. (2022)[3].

**Depression**
Lifetime depression was assessed as part of the mental health follow-up questionnaire (MHQ) using a modified version of the depression module of the Composite International Diagnostic Interview Short Form (CIDI-SF) and defined according to DSM-5 criteria for major depressive disorder[4]. To achieve maximum coverage of the UK Biobank study, we also included in our definition of lifetime depression individuals who had reported during the nurse-led interview at baseline that a doctor had told them that they had depression (UK Biobank data field 20002), participants who reported in response to a single-item question on the MHQ that a professional (doctors, nurse or person with specialist training such as a psychologist or a therapist) had diagnosed them with depression (field 20544), participants with a hospital inpatient record containing an ICD-10 code for depression (F32-F33), participants with a primary care record containing a Read v2 or CTV3 code for depression (see Fabbri et al. (2021)[5] for diagnostic codes and data extraction procedures), and those who were classified as individuals with probable depression according to Smith et al. (2013)[6] based on additional questions that were introduced during the later stages of the baseline assessment.

**Bipolar disorder**
Bipolar disorder was assessed with the MHQ that included several questions on mania and hypomania in combination with the CIDI-SF to assess lifetime depression[4]. We also included in our definition of bipolar disorder individuals who had reported during the nurse-led interview at baseline that a doctor had told them that they had "mania/bipolar disorder/manic depression" (field 20002), participants who responded to a single-item question on the MHQ that a professional had diagnosed them with "mania, hypomania, bipolar or manic-depression" (field 20544), participants with a hospital inpatient record containing an ICD-10 code for mania or bipolar disorder (F30-F31), participants with a primary care record containing a Read v2 or CTV3 code for bipolar disorder (see Fabbri et al. (2021)[5]), and participants who were classified as individuals with probable bipolar disorder according to Smith et al. (2013)[6] based on additional questions that were introduced during the later stages of the baseline assessment.

**Anxiety disorders**
We used a transdiagnostic phenotype for lifetime anxiety disorders and identified cases from multiple sources: the MHQ which included the anxiety disorder module of the CIDI-SF and assessed generalised anxiety disorder according to DSM-5 criteria[4]; individuals with a Generalised Anxiety Disorder Assessment (GAD-7) sum score of ≥10[7], which was assessed as part of the MHQ; individuals who had reported "anxiety/panic attacks" during the nurse-led interview at baseline (field 20002), or "anxiety, nerves or generalized anxiety disorder", "social anxiety or social phobia", "any other phobia", "panic attacks" or "agoraphobia" in response to a single-item question on the MHQ (field 20544); participants with a hospital inpatient record containing an ICD-10 code for anxiety disorders (F40-F41); participants with at least two primary care records containing a Read v2 or CTV3 code for anxiety disorders (see Fabbri et al. (2021)[5]).

#### Frailty phenotype criteria

**Supplementary Table 1.** Frailty phenotype criteria

| Variable | UK Biobank question | UK Biobank data field | Coded in frailty phenotype[1] |
|---|---|---|---|
| Weight loss | "Compared with one year ago, has your weight changed?" | 2306 | Lost weight = 1<br>Other = 0 |
| Exhaustion | "Over the past two weeks, how often have you felt tired or had little energy?" | 120107 | More than half the days = 1<br>Nearly every day = 1<br>Other = 0 |
| Physical activity | "In the last 4 weeks did you spend any time doing the following [activities]?" | 6164[2]<br>1011 (frequency light DIY) | No physical activity = 1<br>Light DIY (≤ 1/week) = 1<br>Light DIY (≥ 2-3/week) = 0<br>Heavy DIY = 0<br>Walking for pleasure = 0<br>Other exercises = 0<br>Strenuous sports = 0 |
| Walking speed | "How would you describe your usual walking pace?" | 924 | Slow = 1<br>Other = 0 |
| Hand-grip strength | Measured during the baseline assessment | 46 (left hand)<br>47 (right hand)<br>1707 (handedness)<br>31 (sex)<br>21001 (BMI) | Maximal grip strength of self-reported dominant hand (adjusted for sex and BMI)[3].<br>Below 20th %tile = 1<br>Equal or above 20th %tile = 0 |

*Note:* BMI = body mass index; DIY = do-it-yourself. [1] "do not know" or "prefer not to answer" were coded as missing data. [2] array variables. [3] grip strength was regressed on sex and BMI, and the residuals from this regression were used to identify individuals in the bottom 20% of the distribution. Criteria adapted from frailty phenotype by Fried et al. (2001), doi: 10.1093/gerona/56.3.m146 and Hanlon et al. (2018), doi: 10.1016/S2468-2667(18)30091-4.

### Frailty index variables

**Supplementary Table 2.** Variables included in the frailty index

| Type of deficit | Item | Variable | UK Biobank data field[1] | Coded in frailty index |
|---|---|---|---|---|
| Sensory | 1 | Glaucoma[2] | 6148[3]<br>2227<br>20002[3] (1277) | Yes = 1<br>No = 0 |
| | 2 | Cataracts[2] | 6148[3]<br>2227<br>20002[3] (1278) | Yes = 1<br>No = 0 |
| | 3 | Hearing difficulty/problems | 2247 | Yes = 1<br>I am completely deaf = 1<br>No = 0 |
| Cranial | 4 | Migraine[2] | 2473<br>20002[3] (1265) | Yes = 1<br>No = 0 |
| | 5 | Mouth/teeth dental problems | 6149 | Any = 1<br>None of the above = 0 |
| Mental wellbeing | 6 | Overall health rating | 2178 | Poor = 1<br>Fair = 0.5<br>Good = 0.25<br>Excellent = 0 |
| | 7 | Frequency of tiredness/lethargy in last two weeks | 2080 | Nearly every day = 1<br>More than half the days = 0.5<br>Several days = 0.25<br>None = 0 |
| | 8 | Sleeplessness/insomnia | 1200 | Usually = 1<br>Sometimes = 0.5<br>Never/rarely = 0 |
| | 9 | Frequency of depressed mood in last two weeks | 2050 | Nearly every day = 1<br>More than half the days = 0.75<br>Several days = 0.5<br>Not at all = 0 |
| | 10 | Nervous feelings | 1970 | Yes = 1<br>No = 0 |
| | 11 | Anxiety / panic attacks[2] | 2473<br>20002[3] (1287) | Yes = 1<br>No = 0 |
| | 12 | Loneliness, isolation | 2020 | Yes = 1<br>No = 0 |
| | 13 | Miserableness | 1930 | Yes = 1<br>No = 0 |
| Infirmity | 14 | Long-standing illness, disability or infirmity | 2188 | Yes = 1<br>No = 0 |
| | 15 | Falls in the last year | 2296 | More than one fall = 1<br>Only one fall = 0.5<br>No falls = 0 |
| | 16 | Fractured/broken bones in last five years | 2463 | Yes = 1<br>No = 0 |
| Cardiometabolic | 17 | Diabetes[2] | 2443<br>20002[3] (1220 & 1223) | Yes = 1<br>No = 0 |
| | 18 | Myocardial infarction[2] | 6150[3]<br>20002[3] (1075) | Yes = 1<br>No = 0 |
| | 19 | Angina[2] | 6150[3]<br>20002[3] (1074) | Yes = 1<br>No = 0 |
| | 20 | Stroke /<br>ischaemic stroke[2] | 6150[3]<br>20002[3] (1081 & 1583) | Yes = 1<br>No = 0 |
| | 21 | Hypertension[2] | 6150[3]<br>20002[3] (1065) | Yes = 1<br>No = 0 |
| | 22 | Hypothyroidism[2] | 2473<br>20002[3] (1226) | Yes = 1<br>No = 0 |
| | 23 | Deep venous thrombosis[2] | 6152[3]<br>20002[3] (1094) | Yes = 1<br>No = 0 |
| | 24 | Cholesterol lowering medication use | 6153 (Females)<br>6177 (Males) | Yes = 1<br>No = 0 |
| Respiratory | 25 | Wheeze or whistling in the chest in last year | 2316 | Yes = 1<br>No = 0 |
| | 26 | Pneumonia[2] | 2473<br>20002[3] (1398) | Yes = 1<br>No = 0 |
| | 27 | Emphysema / chronic bronchitis[2] | 6152[3]<br>20002[3] (1113) | Yes = 1<br>No = 0 |
| | 28 | Asthma[2] | 6152[3] | Yes = 1 |

| | | | | |
|---|---|---|---|---|
| | | | 20002[3] (1111) | No = 0 |
| Musculoskeletal | 29 | Rheumatoid arthritis[2] | 2473<br>20002[3] (1464) | Yes = 1<br>No = 0 |
| | 30 | Osteoarthritis[2] | 2473<br>20002[3] (1465) | Yes = 1<br>No = 0 |
| | 31 | Gout[2] | 2473<br>20002[3] (1466) | Yes = 1<br>No = 0 |
| | 32 | Osteoporosis[2] | 2473<br>20002[3] (1309) | Yes = 1<br>No = 0 |
| Immunological | 33 | Hay fever / allergic rhinitis or eczema/dermatitis[2] | 6152[3]<br>20002[3] (1387 & 1452) | Yes = 1<br>No = 0 |
| | 34 | Psoriasis[2] | 2473<br>20002[3] (1453) | Yes = 1<br>No = 0 |
| Cancer | 35 | Any cancer diagnosis[2] | 134<br>2453 | At least 1 = 1<br>Yes = 1<br>No = 0 |
| | 36 | Multiple cancers diagnosed[2] | 134 | More than 1 = 1<br>0 or 1 = 0 |
| Pain | 37 | Chest pain or discomfort | 2335 | Yes = 1<br>No = 0 |
| | 38 | Headache / neck or shoulder pain (in last month) | 6159[3] | Yes = 1<br>No = 0 |
| | 39 | Back pain (in last month) | 6159[3] | Yes = 1<br>No = 0 |
| | 40 | Stomach or abdominal pain (in last month) | 6159[3] | Yes = 1<br>No = 0 |
| | 41 | Hip pain (in last month) | 6159[3] | Yes = 1<br>No = 0 |
| | 42 | Knee pain (in last month) | 6159[3] | Yes = 1<br>No = 0 |
| | 43 | Pain all over the body (in last month) | 6159[3] | Yes = 1<br>No = 0 |
| | 44 | Facial pain (in last month) | 6159[3] | Yes = 1<br>No = 0 |
| | 45 | Sciatica[2] | 2473<br>20002[3] (1476) | Yes = 1<br>No = 0 |
| Gastrointestinal | 46 | Gastro-oesophageal reflux / gastric reflux[2] | 2473<br>20002[3] (1138) | Yes = 1<br>No = 0 |
| | 47 | Hiatus hernia[2] | 2473<br>20002[3] (1474) | Yes = 1<br>No = 0 |
| | 48 | Cholelithiasis / gall stones[2] | 2473<br>20002[3] (1162) | Yes = 1<br>No = 0 |
| | 49 | Diverticular disease / diverticulitis[2] | 2473<br>20002[3] (1458) | Yes = 1<br>No = 0 |

*Note:* Variables included in frailty index from Williams et al. (2019), doi: 10.1093/gerona/gly094. [1] numbers shown in parentheses are UK Biobank diagnostic codes. [2] conditions diagnosed by a doctor. [3] array variables. For all items with only one corresponding data field, including item 24, "do not know" or "prefer not to answer" responses were coded as missing data. For items with multiple corresponding data fields, we first coded "Yes" and "No" responses using the self-report items and, including for data field 2473, "prefer not to answer" responses as missing data. Additional "Yes" responses were then ascertained from the nurse-led interview diagnostic codes (data field 20002). Individuals with missing data for both data fields 2473 and 20002 were coded as missing data. Items 1 and 2 were coded as "No" if data field 2227 was coded "No" or if data field 6148 was coded "do not know" and data field 20002 was coded "No". Item 17 was coded as "No" if data field 2443 was coded "do not know" and data field 20002 was coded "No". For item 35, "do not know" or "prefer not to answer" responses from data field 2453 were coded as missing data unless participants had at least one cancer for data field 134. For data fields, only data from the first instance (i.e., the baseline assessment) were used.

Overlap between frailty phenotype and frailty index categories

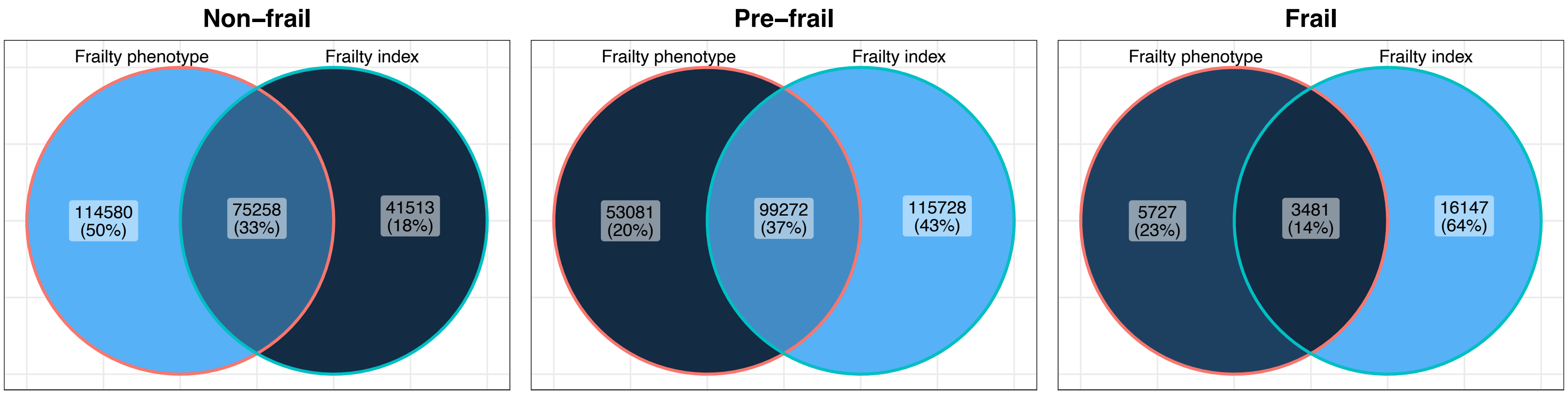

**Supplementary Figure 1.** Venn diagram showing the overlap between individuals with different frailty levels according to the frailty phenotype and frailty index categories. Cut-offs for frailty index categories were non frail (≤ 0.08), pre-frail (>0.08 and <0.25) and frail (≥ 0.25).

#### Sample characteristics stratified by sex

**Supplementary Table 3.** Sample characteristics of individuals with mental disorders and non-psychiatric controls stratified by sex

| | Depression | | Bipolar disorder | | Anxiety disorder | | Controls | |
|---|---|---|---|---|---|---|---|---|
| | Males | Females | Males | Females | Males | Females | Males | Females |
| | N=26546 | N=50040 | N=1319 | N=1710 | N=12835 | N=24944 | N=111347 | N=109447 |
| **Age** | | | | | | | | |
| Mean (SD) | 56.01 (8.04) | 54.67 (7.81) | 55.38 (8.28) | 53.49 (7.68) | 56.43 (7.95) | 55.35 (7.84) | 56.67 (8.27) | 56.19 (8.01) |
| **Neighbourhood deprivation** | | | | | | | | |
| Mean (SD) | -1.48 (2.92) | -1.48 (2.90) | -1.21 (2.99) | -1.01 (3.04) | -1.64 (2.88) | -1.61 (2.86) | -1.78 (2.83) | -1.77 (2.82) |
| **Ethnicity** | | | | | | | | |
| White | 25654 (96.6%) | 48214 (96.4%) | 1247 (94.5%) | 1608 (94.0%) | 12481 (97.2%) | 24273 (97.3%) | 105743 (95.0%) | 103508 (94.6%) |
| Mixed-race | 129 (0.5%) | 393 (0.8%) | 14 (1.1%) | 14 (0.8%) | 52 (0.4%) | 139 (0.6%) | 459 (0.4%) | 646 (0.6%) |
| Black | 148 (0.6%) | 465 (0.9%) | 10 (0.8%) | 31 (1.8%) | 51 (0.4%) | 197 (0.8%) | 1479 (1.3%) | 1801 (1.6%) |
| Asian | 415 (1.6%) | 540 (1.1%) | 30 (2.3%) | 37 (2.2%) | 176 (1.4%) | 175 (0.7%) | 2513 (2.3%) | 2008 (1.8%) |
| Chinese | 31 (0.1%) | 89 (0.2%) | 4 (0.3%) | 2 (0.1%) | 15 (0.1%) | 27 (0.1%) | 322 (0.3%) | 500 (0.5%) |
| Other | 169 (0.6%) | 339 (0.7%) | 14 (1.1%) | 18 (1.1%) | 60 (0.5%) | 133 (0.5%) | 831 (0.7%) | 984 (0.9%) |
| **Highest qualification** | | | | | | | | |
| None | 3836 (14.5%) | 6806 (13.6%) | 147 (11.1%) | 169 (9.9%) | 1737 (13.5%) | 3460 (13.9%) | 17361 (15.6%) | 18308 (16.7%) |
| O levels/GCSEs/CSEs | 6495 (24.5%) | 14814 (29.6%) | 316 (24.0%) | 478 (28.0%) | 3008 (23.4%) | 7457 (29.9%) | 27863 (25.0%) | 33649 (30.7%) |
| A levels/NVQ/HND/HNC[1] | 6346 (23.9%) | 11759 (23.5%) | 308 (23.4%) | 406 (23.7%) | 3087 (24.1%) | 5844 (23.4%) | 27070 (24.3%) | 24198 (22.1%) |
| Degree | 9869 (37.2%) | 16661 (33.3%) | 548 (41.5%) | 657 (38.4%) | 5003 (39.0%) | 8183 (32.8%) | 39053 (35.1%) | 33292 (30.4%) |
| **Spouse/partner cohabitation** | | | | | | | | |
| No | 2418 (9.1%) | 8446 (16.9%) | 149 (11.3%) | 372 (21.8%) | 1041 (8.1%) | 3695 (14.8%) | 6839 (6.1%) | 12309 (11.2%) |
| Yes | 24128 (90.9%) | 41594 (83.1%) | 1170 (88.7%) | 1338 (78.2%) | 11794 (91.9%) | 21249 (85.2%) | 104508 (93.9%) | 97138 (88.8%) |
| **Smoking status** | | | | | | | | |
| Never | 11985 (45.1%) | 27637 (55.2%) | 549 (41.6%) | 859 (50.2%) | 5884 (45.8%) | 13984 (56.1%) | 57470 (51.6%) | 69387 (63.4%) |
| Former | 11075 (41.7%) | 17246 (34.5%) | 534 (40.5%) | 588 (34.4%) | 5424 (42.3%) | 8704 (34.9%) | 42640 (38.3%) | 32725 (29.9%) |
| Current | 3486 (13.1%) | 5157 (10.3%) | 236 (17.9%) | 263 (15.4%) | 1527 (11.9%) | 2256 (9.0%) | 11237 (10.1%) | 7335 (6.7%) |
| **Alcohol intake frequency** | | | | | | | | |
| Never | 1850 (7.0%) | 4392 (8.8%) | 140 (10.6%) | 189 (11.1%) | 882 (6.9%) | 2281 (9.1%) | 5313 (4.8%) | 8998 (8.2%) |
| Special occasions | 2069 (7.8%) | 7418 (14.8%) | 107 (8.1%) | 303 (17.7%) | 890 (6.9%) | 3662 (14.7%) | 6941 (6.2%) | 14413 (13.2%) |
| 1-3/month | 2646 (10.0%) | 6668 (13.3%) | 117 (8.9%) | 235 (13.7%) | 1172 (9.1%) | 3084 (12.4%) | 9269 (8.3%) | 13411 (12.3%) |
| 1-2/week | 6489 (24.4%) | 12612 (25.2%) | 295 (22.4%) | 429 (25.1%) | 3071 (23.9%) | 6200 (24.9%) | 29518 (26.5%) | 29679 (27.1%) |
| 3-4/week | 6600 (24.9%) | 10366 (20.7%) | 314 (23.8%) | 264 (15.4%) | 3256 (25.4%) | 5298 (21.2%) | 31255 (28.1%) | 24668 (22.5%) |
| Daily/almost daily | 6892 (26.0%) | 8584 (17.2%) | 346 (26.2%) | 290 (17.0%) | 3564 (27.8%) | 4419 (17.7%) | 29051 (26.1%) | 18278 (16.7%) |
| **Body mass index** | | | | | | | | |
| Mean (SD) | 28.14 (4.39) | 27.44 (5.38) | 28.51 (4.66) | 27.88 (5.84) | 27.85 (4.27) | 27.10 (5.26) | 27.67 (3.96) | 26.69 (4.83) |
| **Systolic blood pressure** | | | | | | | | |
| Mean (SD) | 139.23 (16.74) | 132.76 (18.47) | 137.58 (16.81) | 130.86 (17.65) | 139.91 (16.84) | 133.81 (18.73) | 141.31 (17.35) | 135.92 (19.43) |
| **Diastolic blood pressure** | | | | | | | | |
| Mean (SD) | 83.78 (9.93) | 80.35 (9.92) | 83.38 (10.08) | 80.10 (9.97) | 83.94 (9.84) | 80.49 (9.90) | 84.18 (9.88) | 80.78 (9.95) |
| **Cholesterol** | | | | | | | | |
| Mean (SD) | 5.49 (1.14) | 5.84 (1.12) | 5.49 (1.14) | 5.81 (1.14) | 5.49 (1.12) | 5.86 (1.12) | 5.53 (1.11) | 5.88 (1.12) |
| **Multimorbidity count** | | | | | | | | |

| | | | | | | | | |
|---|---|---|---|---|---|---|---|---|
| None | 4160 (15.7%) | 8529 (17.0%) | 171 (13.0%) | 209 (12.2%) | 1960 (15.3%) | 3970 (15.9%) | 31250 (28.1%) | 31266 (28.6%) |
| One | 6301 (23.7%) | 11948 (23.9%) | 275 (20.8%) | 376 (22.0%) | 3002 (23.4%) | 5788 (23.2%) | 31513 (28.3%) | 31147 (28.5%) |
| Two | 5474 (20.6%) | 10286 (20.6%) | 272 (20.6%) | 348 (20.4%) | 2721 (21.2%) | 5218 (20.9%) | 22275 (20.0%) | 21283 (19.4%) |
| Three | 4059 (15.3%) | 7597 (15.2%) | 242 (18.3%) | 274 (16.0%) | 2025 (15.8%) | 3909 (15.7%) | 13069 (11.7%) | 12402 (11.3%) |
| Four | 2765 (10.4%) | 4720 (9.4%) | 136 (10.3%) | 206 (12.0%) | 1342 (10.5%) | 2442 (9.8%) | 6800 (6.1%) | 6574 (6.0%) |
| Five or more | 3787 (14.3%) | 6960 (13.9%) | 223 (16.9%) | 297 (17.4%) | 1785 (13.9%) | 3617 (14.5%) | 6440 (5.8%) | 6775 (6.2%) |
| **Antidepressant use** | | | | | | | | |
| No | 21687 (81.7%) | 38639 (77.2%) | 988 (74.9%) | 1187 (69.4%) | 10598 (82.6%) | 19271 (77.3%) | 111347 (100.0%) | 109447 (100.0%) |
| Yes | 4859 (18.3%) | 11401 (22.8%) | 331 (25.1%) | 523 (30.6%) | 2237 (17.4%) | 5673 (22.7%) | 0 (0.0%) | 0 (0.0%) |
| **Antipsychotic use** | | | | | | | | |
| No | 26433 (99.6%) | 49843 (99.6%) | 1245 (94.4%) | 1582 (92.5%) | 12764 (99.4%) | 24822 (99.5%) | 111347 (100.0%) | 109447 (100.0%) |
| Yes | 113 (0.4%) | 197 (0.4%) | 74 (5.6%) | 128 (7.5%) | 71 (0.6%) | 122 (0.5%) | 0 (0.0%) | 0 (0.0%) |
| **Lithium use** | | | | | | | | |
| No | 26514 (99.9%) | 49972 (99.9%) | 1184 (89.8%) | 1571 (91.9%) | 12824 (99.9%) | 24916 (99.9%) | 111347 (100.0%) | 109447 (100.0%) |
| Yes | 32 (0.1%) | 68 (0.1%) | 135 (10.2%) | 139 (8.1%) | 11 (0.1%) | 28 (0.1%) | 0 (0.0%) | 0 (0.0%) |
| **Frailty phenotype** | | | | | | | | |
| Non-frail | 12843 (48.4%) | 22630 (45.2%) | 591 (44.8%) | 693 (40.5%) | 6402 (49.9%) | 11605 (46.5%) | 65107 (58.5%) | 60873 (55.6%) |
| Pre-frail | 12554 (47.3%) | 24995 (50.0%) | 668 (50.6%) | 911 (53.3%) | 5971 (46.5%) | 12228 (49.0%) | 44613 (40.1%) | 46268 (42.3%) |
| Frail | 1149 (4.3%) | 2415 (4.8%) | 60 (4.5%) | 106 (6.2%) | 462 (3.6%) | 1111 (4.5%) | 1627 (1.5%) | 2306 (2.1%) |
| **Frailty phenotype count** | | | | | | | | |
| None | 12843 (48.4%) | 22630 (45.2%) | 591 (44.8%) | 693 (40.5%) | 6402 (49.9%) | 11605 (46.5%) | 65107 (58.5%) | 60873 (55.6%) |
| One | 9294 (35.0%) | 18266 (36.5%) | 493 (37.4%) | 650 (38.0%) | 4544 (35.4%) | 9034 (36.2%) | 36308 (32.6%) | 36371 (33.2%) |
| Two | 3260 (12.3%) | 6729 (13.4%) | 175 (13.3%) | 261 (15.3%) | 1427 (11.1%) | 3194 (12.8%) | 8305 (7.5%) | 9897 (9.0%) |
| Three | 926 (3.5%) | 1944 (3.9%) | 46 (3.5%) | 88 (5.1%) | 372 (2.9%) | 892 (3.6%) | 1398 (1.3%) | 1978 (1.8%) |
| Four | 206 (0.8%) | 426 (0.9%) | 12 (0.9%) | 18 (1.1%) | 82 (0.6%) | 195 (0.8%) | 216 (0.2%) | 307 (0.3%) |
| Five | 17 (0.1%) | 45 (0.1%) | 2 (0.2%) | 0 (0.0%) | 8 (0.1%) | 24 (0.1%) | 13 (0.0%) | 21 (0.0%) |
| Mean (SD) | 0.73 (0.87) | 0.79 (0.89) | 0.79 (0.88) | 0.88 (0.92) | 0.69 (0.84) | 0.76 (0.87) | 0.52 (0.71) | 0.58 (0.76) |
| **Frailty index** | | | | | | | | |
| Mean (SD) | 0.15 (0.08) | 0.15 (0.08) | 0.16 (0.08) | 0.17 (0.08) | 0.15 (0.08) | 0.15 (0.08) | 0.10 (0.06) | 0.11 (0.06) |
| **Frailty index categories** | | | | | | | | |
| Non-frail | 5168 (19.5%) | 9229 (18.4%) | 208 (15.8%) | 237 (13.9%) | 2401 (18.7%) | 4240 (17.0%) | 46028 (41.3%) | 42161 (38.5%) |
| Pre-frail | 18213 (68.6%) | 35067 (70.1%) | 908 (68.8%) | 1190 (69.6%) | 8946 (69.7%) | 17635 (70.7%) | 62127 (55.8%) | 63521 (58.0%) |
| Frail | 3165 (11.9%) | 5744 (11.5%) | 203 (15.4%) | 283 (16.5%) | 1488 (11.6%) | 3069 (12.3%) | 3192 (2.9%) | 3765 (3.4%) |

*Note:* SD = standard deviation; GCSEs = general certificate of secondary education; CSE = certificate of secondary education; NVQ = national vocational qualification; HND = higher national diploma; HNC = higher national certificate. [1] also includes 'other professional qualifications'. Cut-offs for frailty index categories were non frail (≤ 0.08), pre-frail (>0.08 and <0.25) and frail (≥ 0.25).

Case-control differences stratified by sex

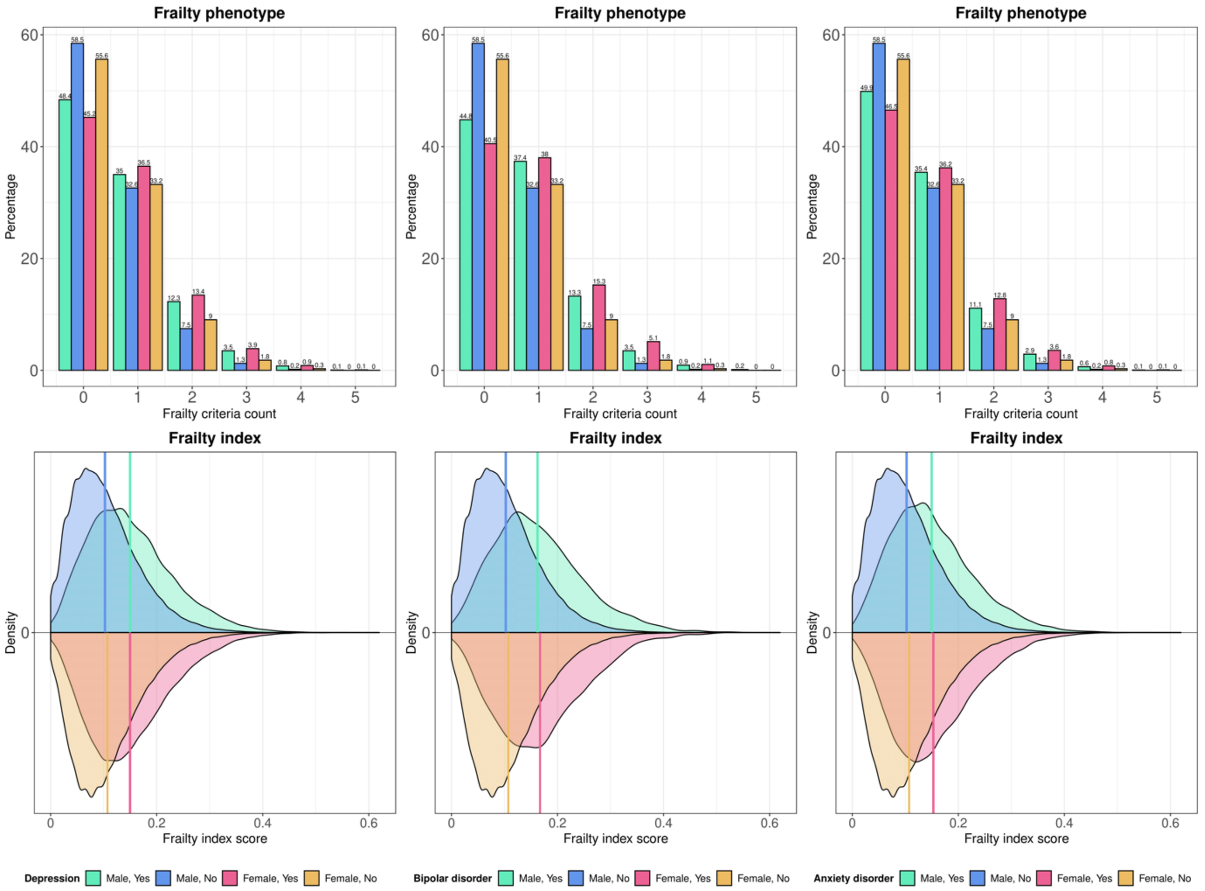

**Supplementary Figure 1.** Histograms and density plots showing the distribution of the frailty phenotype criteria count (top panels) and the frailty index (bottom panels), respectively, for individuals with mental disorders and non-psychiatric controls stratified by sex.

#### Sex differences in frailty index

**Supplementary Table 4.** Sex differences in the frailty index

| Term | SMD | 95% CI | | $p_{Bonf.}$ | $p_{BH}$ |
|---|---|---|---|---|---|
| Controls | 0.072 | 0.064 | 0.081 | <0.001 | <0.001 |
| Depression | 0.001 | -0.014 | 0.016 | >0.999 | 0.891 |
| Bipolar disorder | 0.055 | -0.017 | 0.127 | 0.536 | 0.179 |
| Anxiety disorder | 0.039 | 0.017 | 0.060 | 0.002 | <0.001 |

*Note:* SMD = standardised mean difference; CI = confidence interval; Bonf. = Bonferroni; BH = Benjamini & Hochberg.

#### Sex-stratified regression models

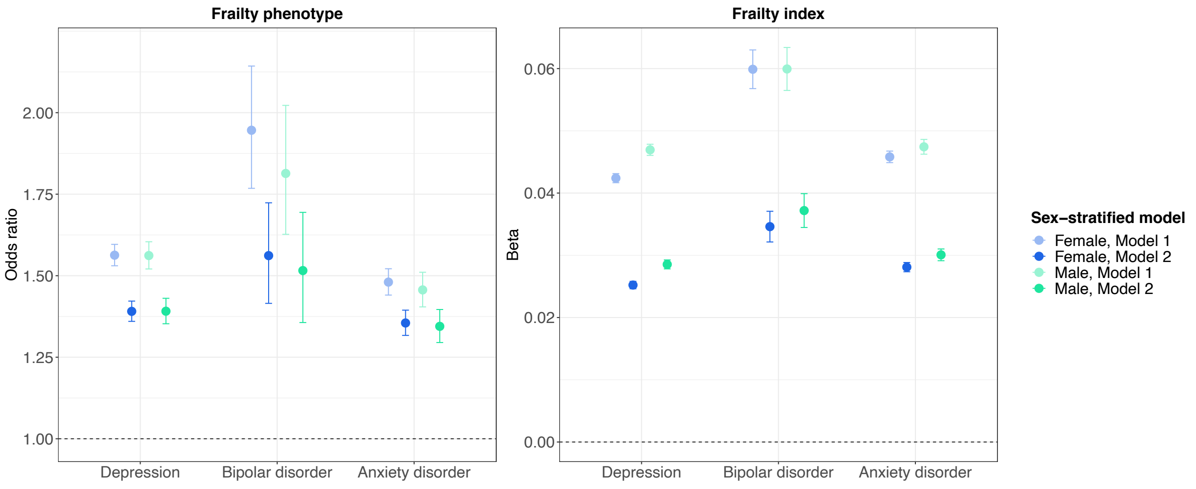

**Supplementary Figure 2.** Frailty in individuals with mental disorders compared to non-psychiatric controls (reference group) stratified by sex. Estimates shown for the frailty phenotype are odds ratios and 95% confidence intervals (CI) from ordinal logistic regression models, indicating changes in odds of being frailer associated with being in the case group relative to the control group. Estimates shown for the frailty index are ordinary least squares regression beta coefficients and 95% CI. Model 1 – unadjusted; Model 2 – adjusted for age, sex, ethnicity, highest qualification, Townsend deprivation index, cohabitation with spouse/partner, smoking status, alcohol intake frequency, systolic and diastolic blood pressure, body mass index, cholesterol, multimorbidity count and assessment centre.

**Supplementary Table 5.** Frailty in males and females with mental disorders and non-psychiatric controls

| | **Males** | | | | | | **Females** | | | | | |
|---|---|---|---|---|---|---|---|---|---|---|---|---|
| | Model 1 | | | Model 2 | | | Model 1 | | | Model 2 | | |
| **Frailty phenotype** | OR | 95% CI | | *β* | 95% CI | | OR | 95% CI | | *β* | 95% CI | |
| Controls | Ref | - | - | Ref | - | - | Ref | - | - | Ref | - | - |
| Depression | 1.562 | 1.521 | 1.604 | 1.391 | 1.353 | 1.431 | 1.563 | 1.531 | 1.596 | 1.391 | 1.360 | 1.422 |
| Bipolar disorder | 1.814 | 1.627 | 2.022 | 1.516 | 1.356 | 1.694 | 1.946 | 1.768 | 2.143 | 1.562 | 1.415 | 1.724 |
| Anxiety disorder | 1.457 | 1.404 | 1.511 | 1.345 | 1.295 | 1.396 | 1.480 | 1.441 | 1.521 | 1.355 | 1.317 | 1.394 |
| **Frailty index** | *β* | 95% CI | | OR | 95% CI | | *β* | 95% CI | | OR | 95% CI | |
| Controls | Ref | - | - | Ref | - | - | Ref | - | - | Ref | - | - |
| Depression | 0.047 | 0.046 | 0.048 | 0.029 | 0.028 | 0.029 | 0.042 | 0.042 | 0.043 | 0.025 | 0.025 | 0.026 |
| Bipolar disorder | 0.060 | 0.056 | 0.063 | 0.037 | 0.034 | 0.040 | 0.060 | 0.057 | 0.063 | 0.035 | 0.032 | 0.037 |
| Anxiety disorder | 0.047 | 0.046 | 0.049 | 0.030 | 0.029 | 0.031 | 0.046 | 0.045 | 0.047 | 0.028 | 0.027 | 0.029 |

*Note:* OR = odds ratio; *β* = ordinary least squares regression beta coefficient; CI = confidence interval; Ref = reference group. All Bonferroni-adjusted *p*-values < 0.001. Odds ratios indicate changes in odds of being frailer associated with being in the case group relative to the control group. Model 1 – unadjusted; Model 2 – adjusted for age, sex, ethnicity, highest qualification, Townsend deprivation index, cohabitation with spouse/partner, smoking status, alcohol intake frequency, systolic and diastolic blood pressure, body mass index, cholesterol, multimorbidity count and assessment centre.

#### Frailty phenotype by age

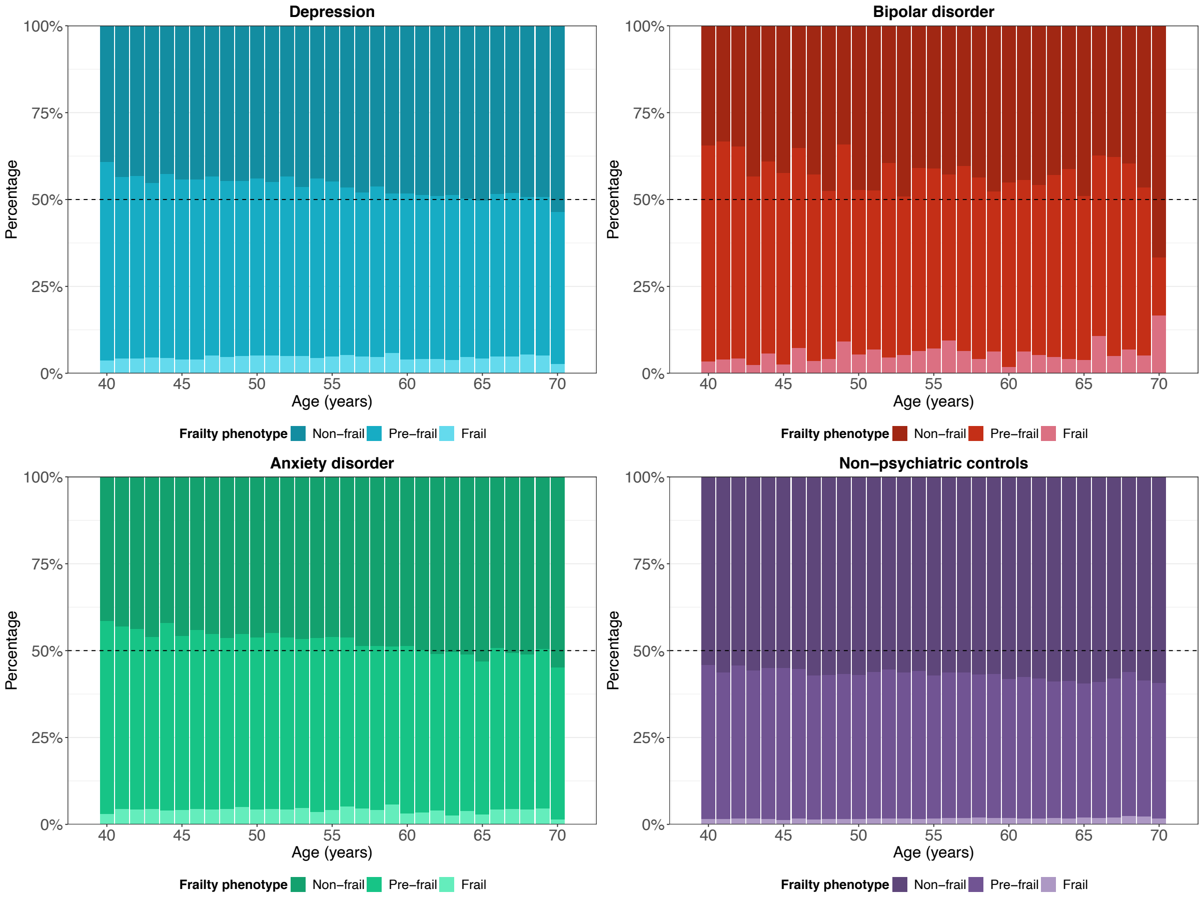

**Supplementary Figure 3.** Stacked bar charts showing the frailty phenotype by age in individuals with mental disorders and non-psychiatric controls.

### Descriptive statistics follow-up

**Supplementary Table 6.** Descriptive statistics follow-up

| | | **Depression** | | **Bipolar disorder** | | **Anxiety disorder** | |
|---|---|---|---|---|---|---|---|
| Full sample size (cases) | | 297380 (76586) | | 223823 (3029) | | 258573 (37779) | |
| Person-years follow-up | | 3516706 | | 2654566 | | 3066828 | |
| Median (IQR) censored | | 12.09 (1.35) | | 12.19 (1.31) | | 12.14 (1.31) | |
| Deaths, all (cases) | | 17240 (4138) | | 13315 (213) | | 15052 (1950) | |
| **Frailty phenotype** | | Censored | Deaths | Censored | Deaths | Censored | Deaths |
| Non-frail | No | 119473 | 6507 | 119473 | 6507 | 119473 | 6507 |
| Non-frail | Yes | 33898 | 1575 | 1209 | 75 | 17184 | 823 |
| Pre-frail | No | 84865 | 6016 | 84865 | 6016 | 84865 | 6016 |
| Pre-frail | Yes | 35438 | 2111 | 1463 | 116 | 17264 | 935 |
| Frail | No | 3354 | 579 | 3354 | 579 | 3354 | 579 |
| Frail | Yes | 3112 | 452 | 144 | 22 | 1381 | 192 |
| **Frailty index categories** | | | | | | | |
| Non-frail | No | 84753 | 3436 | 84753 | 3436 | 84753 | 3436 |
| Non-frail | Yes | 13927 | 470 | 428 | 17 | 6408 | 233 |
| Pre-frail | No | 117045 | 8603 | 117045 | 8603 | 117045 | 8603 |
| Pre-frail | Yes | 50584 | 2696 | 1960 | 138 | 25289 | 1292 |
| Frail | No | 5894 | 1063 | 5894 | 1063 | 5894 | 1063 |
| Frail | Yes | 7937 | 972 | 428 | 58 | 4132 | 425 |

*Note:* IQR = interquartile range.

All-cause mortality by case status

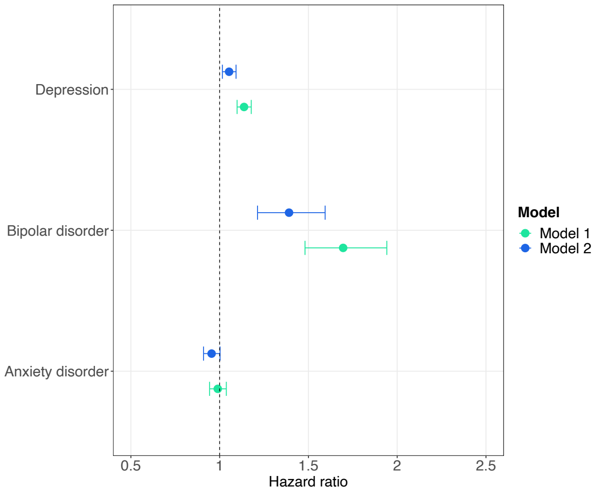

**Supplementary Figure 4.** All-cause mortality in individuals with mental disorders and non-psychiatric controls (reference group). Hazard ratios (HR) with 95% confidence intervals from Cox proportional hazards models. Age (in years) was used as the underlying time axis. Model 1 – unadjusted; Model 2 – adjusted for sex, ethnicity, highest qualification, Townsend deprivation index, cohabitation with spouse/partner, smoking status, alcohol intake frequency, systolic and diastolic blood pressure, body mass index, cholesterol, multimorbidity count and assessment centre.

All-cause mortality by frailty level

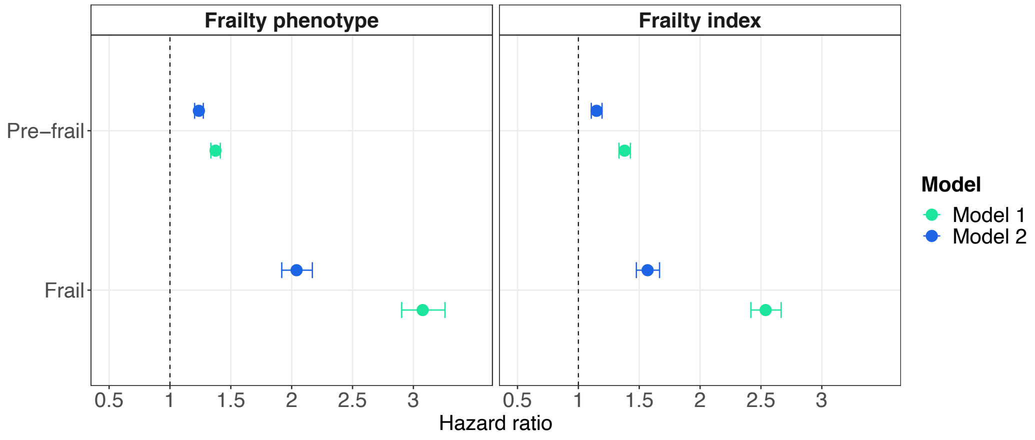

**Supplementary Figure 5.** All-cause mortality by frailty level (non-frail, pre-frail and frail). Reference group: non-frail participants. Hazard ratios (HR) with 95% confidence intervals from Cox proportional hazards models. Age (in years) was used as the underlying time axis. Model 1 – unadjusted; Model 2 – adjusted for sex, ethnicity, highest qualification, Townsend deprivation index, cohabitation with spouse/partner, smoking status, alcohol intake frequency, systolic and diastolic blood pressure, body mass index, cholesterol, multimorbidity count and assessment centre.

Survival probabilities by frailty phenotype and case status

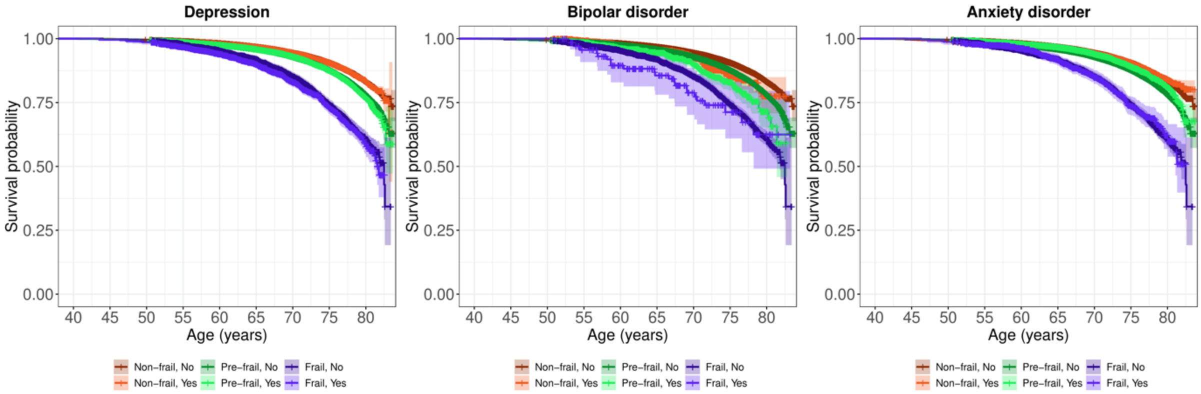

**Supplementary Figure 6.** Kaplan-Meier survival probabilities for all-cause mortality by frailty phenotype (non-frail, pre-frail and frail) in individuals with mental disorders (yes) and non-psychiatric controls (no). Two observations (one left panel and one right panel) were removed due to the death occurring after the maximum censoring age. All log-rank test $p$-values $< 0.001$.

Survival probabilities by frailty index categories and case status

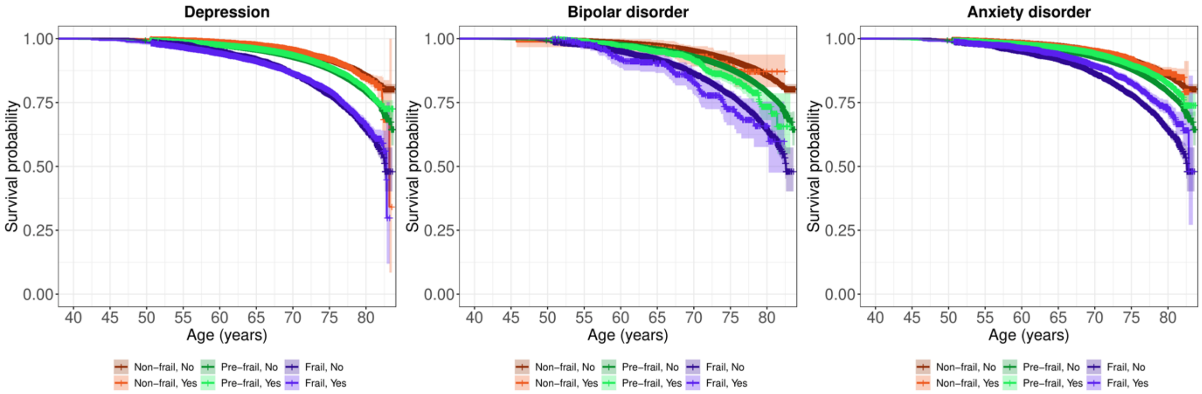

**Supplementary Figure 7.** Kaplan-Meier survival probabilities for all-cause mortality by frailty index categories (non-frail, pre-frail and frail) in individuals with mental disorders (yes) and non-psychiatric controls (no). All log-rank test *p*-values $< 0.001$.

All-cause mortality by frailty phenotype and case status

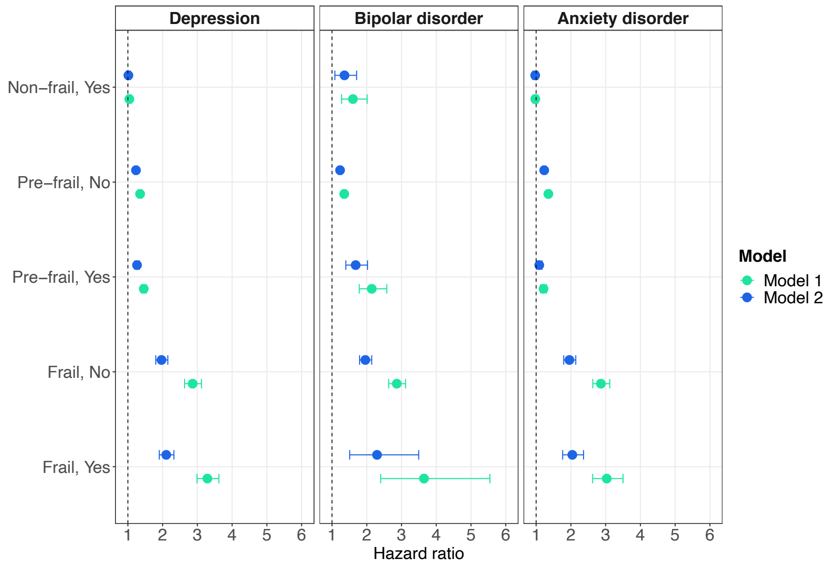

**Supplementary Figure 8.** All-cause mortality by frailty phenotype (non-frail, pre-frail and frail) in individuals with mental disorders and non-psychiatric controls. Reference group: non-frail controls. Hazard ratios (HR) with 95% confidence intervals from Cox proportional hazards models. Age (in years) was used as the underlying time axis. Model 1 – unadjusted; Model 2 – adjusted for sex, ethnicity, highest qualification, Townsend deprivation index, cohabitation with spouse/partner, smoking status, alcohol intake frequency, systolic and diastolic blood pressure, body mass index, cholesterol, multimorbidity count and assessment centre.

All-cause mortality by frailty index categories and case status

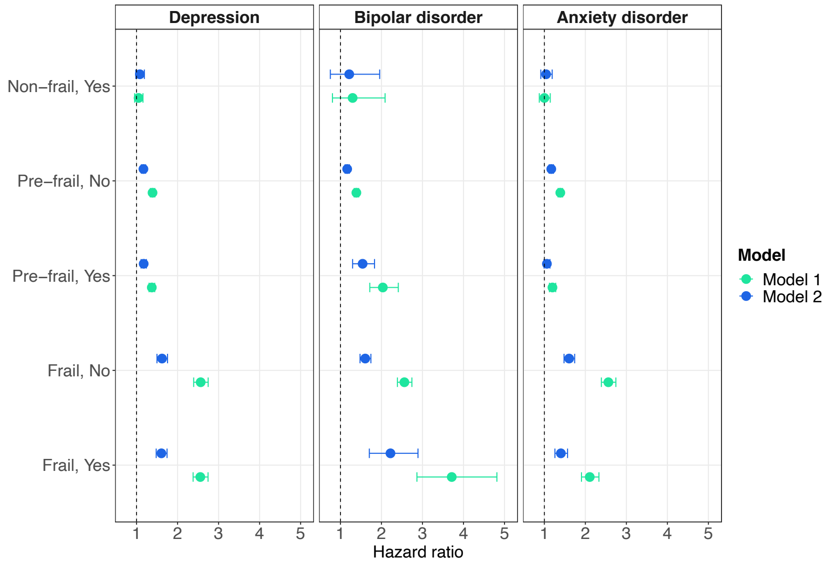

**Supplementary Figure 9.** All-cause mortality by frailty index categories in individuals with mental disorders and non-psychiatric controls. Reference group: non-frail controls. Hazard ratios (HR) with 95% confidence intervals from Cox proportional hazards models. Age (in years) was used as the underlying time axis. Model 1 – unadjusted; Model 2 – adjusted for sex, ethnicity, highest qualification, Townsend deprivation index, cohabitation with spouse/partner, smoking status, alcohol intake frequency, systolic and diastolic blood pressure, body mass index, cholesterol, multimorbidity count and assessment centre.

All-cause mortality by frailty phenotype and case status stratified by sex

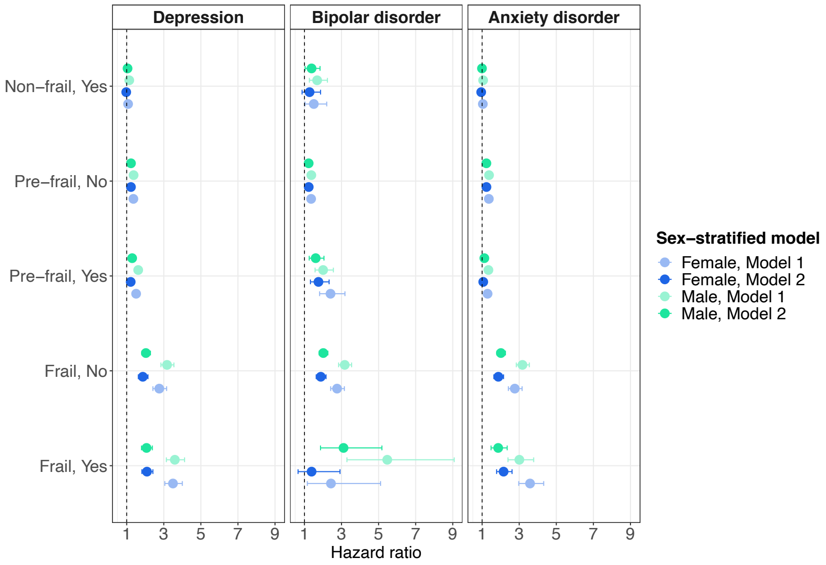

**Supplementary Figure 10.** All-cause mortality by frailty phenotype (non-frail, pre-frail and frail) in individuals with mental disorders and non-psychiatric controls stratified by sex. Reference group: non-frail controls. Hazard ratios (HR) with 95% confidence intervals from Cox proportional hazards models. Age (in years) was used as the underlying time axis. Model 1 – unadjusted; Model 2 – adjusted for ethnicity, highest qualification, Townsend deprivation index, cohabitation with spouse/partner, smoking status, alcohol intake frequency, systolic and diastolic blood pressure, body mass index, cholesterol, multimorbidity count and assessment centre.

**Supplementary Table 7.** All-cause mortality by frailty phenotype in individuals with mental disorders and non-psychiatric controls stratified by sex

| | | **Males** | | | | | | | | | | **Females** | | | | | | | | | |
|---|---|---|---|---|---|---|---|---|---|---|---|---|---|---|---|---|---|---|---|---|---|
| | | **Model 1** | | | | | **Model 2** | | | | | **Model 1** | | | | | **Model 2** | | | | |
| **Depression** | | HR | 95% CI | | $p_{Bonf.}$ | $p_{BH}$ | HR | 95% CI | | $p_{Bonf.}$ | $p_{BH}$ | HR | 95% CI | | $p_{Bonf.}$ | $p_{BH}$ | HR | 95% CI | | $p_{Bonf.}$ | $p_{BH}$ |
| Non-frail | No | Ref | - | - | - | - | Ref | - | - | - | - | Ref | - | - | - | - | Ref | - | - | - | - |
| Non-frail | Yes | 1.139 | 1.057 | 1.228 | 0.019 | <0.001 | 1.046 | 0.970 | 1.129 | >0.999 | 0.256 | 1.076 | 0.991 | 1.168 | >0.999 | 0.104 | 0.975 | 0.897 | 1.060 | >0.999 | 0.555 |
| Pre-frail | No | 1.372 | 1.313 | 1.434 | <0.001 | <0.001 | 1.232 | 1.178 | 1.288 | <0.001 | <0.001 | 1.361 | 1.285 | 1.443 | <0.001 | <0.001 | 1.230 | 1.160 | 1.304 | <0.001 | <0.001 |
| Pre-frail | Yes | 1.621 | 1.516 | 1.734 | <0.001 | <0.001 | 1.295 | 1.208 | 1.388 | <0.001 | <0.001 | 1.509 | 1.402 | 1.623 | <0.001 | <0.001 | 1.216 | 1.127 | 1.312 | <0.001 | <0.001 |
| Frail | No | 3.176 | 2.845 | 3.546 | <0.001 | <0.001 | 2.036 | 1.817 | 2.282 | <0.001 | <0.001 | 2.759 | 2.413 | 3.153 | <0.001 | <0.001 | 1.870 | 1.628 | 2.148 | <0.001 | <0.001 |
| Frail | Yes | 3.597 | 3.139 | 4.122 | <0.001 | <0.001 | 2.070 | 1.797 | 2.384 | <0.001 | <0.001 | 3.496 | 3.055 | 4.001 | <0.001 | <0.001 | 2.094 | 1.817 | 2.413 | <0.001 | <0.001 |
| **Bipolar disorder** | | HR | 95% CI | | $p_{Bonf.}$ | $p_{BH}$ | HR | 95% CI | | $p_{Bonf.}$ | $p_{BH}$ | HR | 95% CI | | $p_{Bonf.}$ | $p_{BH}$ | HR | 95% CI | | $p_{Bonf.}$ | $p_{BH}$ |
| Non-frail | No | Ref | - | - | - | - | Ref | - | - | - | - | Ref | - | - | - | - | Ref | - | - | - | - |
| Non-frail | Yes | 1.686 | 1.268 | 2.241 | 0.010 | <0.001 | 1.385 | 1.041 | 1.843 | 0.758 | 0.028 | 1.507 | 1.031 | 2.203 | >0.999 | 0.044 | 1.278 | 0.874 | 1.870 | >0.999 | 0.247 |
| Pre-frail | No | 1.372 | 1.313 | 1.434 | <0.001 | <0.001 | 1.230 | 1.176 | 1.286 | <0.001 | <0.001 | 1.362 | 1.285 | 1.443 | <0.001 | <0.001 | 1.233 | 1.162 | 1.308 | <0.001 | <0.001 |
| Pre-frail | Yes | 2.009 | 1.575 | 2.562 | <0.001 | <0.001 | 1.609 | 1.260 | 2.054 | 0.004 | <0.001 | 2.408 | 1.819 | 3.186 | <0.001 | <0.001 | 1.758 | 1.325 | 2.332 | 0.003 | <0.001 |
| Frail | No | 3.175 | 2.844 | 3.545 | <0.001 | <0.001 | 2.018 | 1.799 | 2.264 | <0.001 | <0.001 | 2.761 | 2.415 | 3.156 | <0.001 | <0.001 | 1.883 | 1.636 | 2.167 | <0.001 | <0.001 |
| Frail | Yes | 5.468 | 3.294 | 9.079 | <0.001 | <0.001 | 3.110 | 1.868 | 5.178 | <0.001 | <0.001 | 2.431 | 1.158 | 5.107 | 0.568 | 0.026 | 1.386 | 0.658 | 2.919 | >0.999 | 0.418 |
| **Anxiety disorder** | | HR | 95% CI | | $p_{Bonf.}$ | $p_{BH}$ | HR | 95% CI | | $p_{Bonf.}$ | $p_{BH}$ | HR | 95% CI | | $p_{Bonf.}$ | $p_{BH}$ | HR | 95% CI | | $p_{Bonf.}$ | $p_{BH}$ |
| Non-frail | No | Ref | - | - | - | - | Ref | - | - | - | - | Ref | - | - | - | - | Ref | - | - | - | - |
| Non-frail | Yes | 1.053 | 0.952 | 1.165 | >0.999 | 0.326 | 0.991 | 0.895 | 1.097 | >0.999 | 0.863 | 1.038 | 0.935 | 1.153 | >0.999 | 0.501 | 0.949 | 0.854 | 1.055 | >0.999 | 0.372 |
| Pre-frail | No | 1.372 | 1.313 | 1.434 | <0.001 | <0.001 | 1.229 | 1.175 | 1.286 | <0.001 | <0.001 | 1.362 | 1.285 | 1.443 | <0.001 | <0.001 | 1.232 | 1.161 | 1.307 | <0.001 | <0.001 |
| Pre-frail | Yes | 1.339 | 1.214 | 1.476 | <0.001 | <0.001 | 1.119 | 1.014 | 1.236 | 0.767 | 0.028 | 1.293 | 1.173 | 1.426 | <0.001 | <0.001 | 1.060 | 0.960 | 1.172 | >0.999 | 0.288 |
| Frail | No | 3.175 | 2.844 | 3.545 | <0.001 | <0.001 | 2.012 | 1.794 | 2.256 | <0.001 | <0.001 | 2.761 | 2.415 | 3.156 | <0.001 | <0.001 | 1.875 | 1.631 | 2.156 | <0.001 | <0.001 |
| Frail | Yes | 3.008 | 2.394 | 3.779 | <0.001 | <0.001 | 1.866 | 1.481 | 2.352 | <0.001 | <0.001 | 3.584 | 2.974 | 4.319 | <0.001 | <0.001 | 2.157 | 1.780 | 2.614 | <0.001 | <0.001 |

*Note:* HR = hazard ratio; CI = confidence interval; Bonf. = Bonferroni; BH = Benjamini & Hochberg. Age (in years) was used as the underlying time axis. Model 1 – unadjusted; Model 2 – adjusted for ethnicity, highest qualification, Townsend deprivation index, cohabitation with spouse/partner, smoking status, alcohol intake frequency, systolic and diastolic blood pressure, body mass index, cholesterol, multimorbidity count and assessment centre.

All-cause mortality by frailty index categories and case status stratified by sex

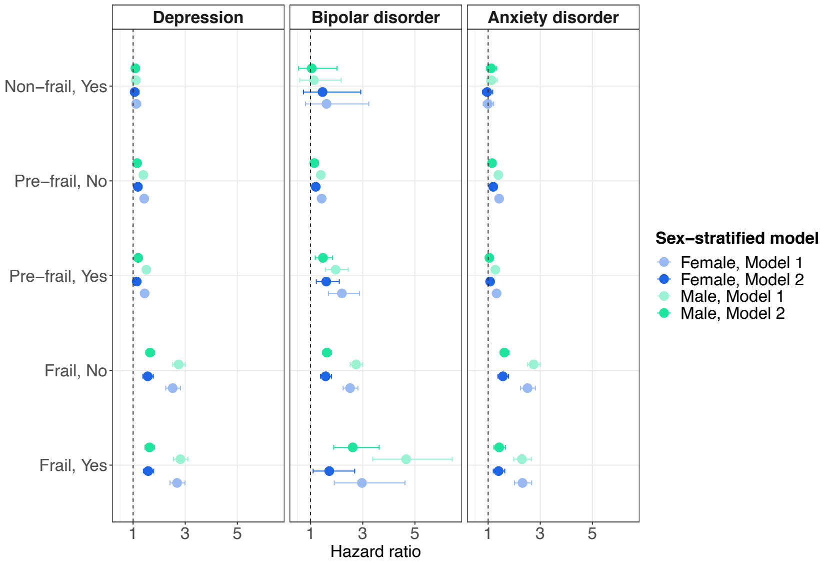

**Supplementary Figure 11.** All-cause mortality by frailty index categories (non-frail, pre-frail and frail) in individuals with mental disorders and non-psychiatric controls stratified by sex. Reference group: non-frail controls. Hazard ratios (HR) with 95% confidence intervals from Cox proportional hazards models. Age (in years) was used as the underlying time axis. Model 1 – unadjusted; Model 2 – adjusted for ethnicity, highest qualification, Townsend deprivation index, cohabitation with spouse/partner, smoking status, alcohol intake frequency, systolic and diastolic blood pressure, body mass index, cholesterol, multimorbidity count and assessment centre.

**Supplementary Table 8.** All-cause mortality by frailty index categories in individuals with mental disorders and non-psychiatric controls stratified by sex

| | | **Males** | | | | | | | | | | **Females** | | | | | | | | | |
|---|---|---|---|---|---|---|---|---|---|---|---|---|---|---|---|---|---|---|---|---|---|
| | | **Model 1** | | | | | **Model 2** | | | | | **Model 1** | | | | | **Model 2** | | | | |
| **Depression** | | HR | 95% CI | | $p_{Bonf.}$ | $p_{BH}$ | HR | 95% CI | | $p_{Bonf.}$ | $p_{BH}$ | HR | 95% CI | | $p_{Bonf.}$ | $p_{BH}$ | HR | 95% CI | | $p_{Bonf.}$ | $p_{BH}$ |
| Non-frail | No | Ref | - | - | - | - | Ref | - | - | - | - | Ref | - | - | - | - | Ref | - | - | - | - |
| Non-frail | Yes | 1.107 | 0.970 | 1.264 | >0.999 | 0.165 | 1.085 | 0.950 | 1.239 | >0.999 | 0.259 | 1.120 | 0.972 | 1.291 | >0.999 | 0.146 | 1.062 | 0.921 | 1.224 | >0.999 | 0.440 |
| Pre-frail | No | 1.393 | 1.326 | 1.464 | <0.001 | <0.001 | 1.155 | 1.093 | 1.221 | <0.001 | <0.001 | 1.425 | 1.333 | 1.522 | <0.001 | <0.001 | 1.185 | 1.102 | 1.275 | <0.001 | <0.001 |
| Pre-frail | Yes | 1.508 | 1.410 | 1.613 | <0.001 | <0.001 | 1.197 | 1.113 | 1.288 | <0.001 | <0.001 | 1.443 | 1.334 | 1.560 | <0.001 | <0.001 | 1.140 | 1.047 | 1.242 | 0.076 | 0.003 |
| Frail | No | 2.747 | 2.517 | 2.998 | <0.001 | <0.001 | 1.649 | 1.490 | 1.825 | <0.001 | <0.001 | 2.520 | 2.253 | 2.818 | <0.001 | <0.001 | 1.561 | 1.374 | 1.773 | <0.001 | <0.001 |
| Frail | Yes | 2.813 | 2.552 | 3.101 | <0.001 | <0.001 | 1.629 | 1.458 | 1.821 | <0.001 | <0.001 | 2.688 | 2.417 | 2.989 | <0.001 | <0.001 | 1.581 | 1.396 | 1.789 | <0.001 | <0.001 |
| **Bipolar disorder** | | HR | 95% CI | | $p_{Bonf.}$ | $p_{BH}$ | HR | 95% CI | | $p_{Bonf.}$ | $p_{BH}$ | HR | 95% CI | | $p_{Bonf.}$ | $p_{BH}$ | HR | 95% CI | | $p_{Bonf.}$ | $p_{BH}$ |
| Non-frail | No | Ref | - | - | - | - | Ref | - | - | - | - | Ref | - | - | - | - | Ref | - | - | - | - |
| Non-frail | Yes | 1.127 | 0.586 | 2.170 | >0.999 | 0.744 | 1.045 | 0.543 | 2.012 | >0.999 | 0.896 | 1.612 | 0.804 | 3.230 | >0.999 | 0.206 | 1.457 | 0.727 | 2.924 | >0.999 | 0.321 |
| Pre-frail | No | 1.392 | 1.325 | 1.463 | <0.001 | <0.001 | 1.144 | 1.081 | 1.211 | <0.001 | <0.001 | 1.423 | 1.331 | 1.521 | <0.001 | <0.001 | 1.198 | 1.112 | 1.290 | <0.001 | <0.001 |
| Pre-frail | Yes | 1.959 | 1.572 | 2.442 | <0.001 | <0.001 | 1.474 | 1.179 | 1.842 | 0.020 | <0.001 | 2.201 | 1.684 | 2.878 | <0.001 | <0.001 | 1.601 | 1.220 | 2.101 | 0.021 | <0.001 |
| Frail | No | 2.745 | 2.515 | 2.995 | <0.001 | <0.001 | 1.624 | 1.463 | 1.802 | <0.001 | <0.001 | 2.515 | 2.248 | 2.813 | <0.001 | <0.001 | 1.577 | 1.380 | 1.803 | <0.001 | <0.001 |
| Frail | Yes | 4.664 | 3.384 | 6.427 | <0.001 | <0.001 | 2.616 | 1.886 | 3.628 | <0.001 | <0.001 | 2.969 | 1.909 | 4.619 | <0.001 | <0.001 | 1.717 | 1.096 | 2.690 | 0.546 | 0.024 |
| **Anxiety disorder** | | HR | 95% CI | | $p_{Bonf.}$ | $p_{BH}$ | HR | 95% CI | | $p_{Bonf.}$ | $p_{BH}$ | HR | 95% CI | | $p_{Bonf.}$ | $p_{BH}$ | HR | 95% CI | | $p_{Bonf.}$ | $p_{BH}$ |
| Non-frail | No | Ref | - | - | - | - | Ref | - | - | - | - | Ref | - | - | - | - | Ref | - | - | - | - |
| Non-frail | Yes | 1.129 | 0.945 | 1.348 | >0.999 | 0.218 | 1.114 | 0.933 | 1.331 | >0.999 | 0.259 | 0.992 | 0.812 | 1.212 | >0.999 | 0.940 | 0.962 | 0.787 | 1.176 | >0.999 | 0.728 |
| Pre-frail | No | 1.393 | 1.326 | 1.464 | <0.001 | <0.001 | 1.151 | 1.088 | 1.217 | <0.001 | <0.001 | 1.422 | 1.331 | 1.520 | <0.001 | <0.001 | 1.197 | 1.112 | 1.289 | <0.001 | <0.001 |
| Pre-frail | Yes | 1.275 | 1.165 | 1.394 | <0.001 | <0.001 | 1.047 | 0.953 | 1.151 | >0.999 | 0.360 | 1.325 | 1.206 | 1.456 | <0.001 | <0.001 | 1.078 | 0.975 | 1.192 | >0.999 | 0.173 |
| Frail | No | 2.747 | 2.517 | 2.998 | <0.001 | <0.001 | 1.623 | 1.464 | 1.799 | <0.001 | <0.001 | 2.513 | 2.247 | 2.810 | <0.001 | <0.001 | 1.566 | 1.375 | 1.784 | <0.001 | <0.001 |
| Frail | Yes | 2.296 | 1.983 | 2.658 | <0.001 | <0.001 | 1.425 | 1.220 | 1.666 | <0.001 | <0.001 | 2.318 | 2.013 | 2.670 | <0.001 | <0.001 | 1.401 | 1.198 | 1.639 | <0.001 | <0.001 |

*Note:* HR = hazard ratio; CI = confidence interval; Bonf. = Bonferroni; BH = Benjamini & Hochberg. Age (in years) was used as the underlying time axis. Model 1 – unadjusted; Model 2 – adjusted for ethnicity, highest qualification, Townsend deprivation index, cohabitation with spouse/partner, smoking status, alcohol intake frequency, systolic and diastolic blood pressure, body mass index, cholesterol, multimorbidity count and assessment centre.

### Sensitivity analysis

#### Excluding individuals with comorbid depression and anxiety disorders

**Supplementary Table 9.** Descriptive statistics follow-up, no comorbid depression and anxiety disorders

| | | **Depression** | | **Anxiety disorder** | |
|---|---|---|---|---|---|
| Full sample size (cases) | | 283165 (48737) | | 248495 (14067) | |
| Person-years follow-up | | 3351539 | | 2950320 | |
| Median (IQR) censored | | 12.10 (1.35) | | 12.19 (1.31) | |
| Deaths, all (cases) | | 16236 (2823) | | 14261 (848) | |
| **Frailty phenotype** | | Censored | Deaths | Censored | Deaths |
| Non-frail | No | 127228 | 6666 | 127228 | 6666 |
| Non-frail | Yes | 21989 | 1055 | 7064 | 386 |
| Pre-frail | No | 90285 | 6160 | 90285 | 6160 |
| Pre-frail | Yes | 22063 | 1472 | 5854 | 397 |
| Frail | No | 3502 | 587 | 3502 | 587 |
| Frail | Yes | 1862 | 296 | 301 | 65 |
| **Frailty index categories** | | | | | |
| Non-frail | No | 89247 | 3513 | 89247 | 3513 |
| Non-frail | Yes | 10097 | 349 | 3162 | 129 |
| Pre-frail | No | 125561 | 8822 | 125561 | 8822 |
| Pre-frail | Yes | 31569 | 1870 | 9133 | 599 |
| Frail | No | 6207 | 1078 | 6207 | 1078 |
| Frail | Yes | 4248 | 604 | 924 | 120 |

*Note:* IQR = interquartile range.

**Supplementary Table 10.** All-cause mortality by frailty phenotype, no comorbid depression and anxiety disorders

| | | Model 1 | | | | | | Model 2 | | | | | |
|---|---|---|---|---|---|---|---|---|---|---|---|---|---|
| **Depression** | | HR | 95% CI | | $p_{Bonf.}$ | $p_{BH}$ | RD % | HR | 95% CI | | $p_{Bonf.}$ | $p_{BH}$ | RD % |
| Non-frail | No | Ref | - | - | - | - | - | Ref | - | - | - | - | - |
| Non-frail | Yes | 1.101 | 1.032 | 1.175 | 0.076 | 0.004 | - | 1.069 | 1.001 | 1.141 | 0.945 | 0.053 | - |
| Pre-frail | No | 1.353 | 1.307 | 1.401 | <0.001 | <0.001 | - | 1.236 | 1.193 | 1.280 | <0.001 | <0.001 | - |
| Pre-frail | Yes | 1.640 | 1.550 | 1.735 | <0.001 | <0.001 | 81.23 | 1.413 | 1.333 | 1.497 | <0.001 | <0.001 | 74.89 |
| Frail | No | 2.885 | 2.651 | 3.139 | <0.001 | <0.001 | - | 1.983 | 1.817 | 2.164 | <0.001 | <0.001 | - |
| Frail | Yes | 3.557 | 3.166 | 3.996 | <0.001 | <0.001 | 35.68 | 2.212 | 1.961 | 2.494 | <0.001 | <0.001 | 23.29 |
| **Anxiety disorder** | | | | | | | | | | | | | |
| Non-frail | No | Ref | - | - | - | - | - | Ref | - | - | - | - | - |
| Non-frail | Yes | 0.992 | 0.896 | 1.099 | >0.999 | 0.883 | - | 1.011 | 0.912 | 1.121 | >0.999 | 0.875 | - |
| Pre-frail | No | 1.353 | 1.307 | 1.401 | <0.001 | <0.001 | - | 1.234 | 1.191 | 1.278 | <0.001 | <0.001 | - |
| Pre-frail | Yes | 1.263 | 1.141 | 1.398 | <0.001 | <0.001 | -25.56 | 1.175 | 1.061 | 1.300 | 0.039 | 0.003 | -25.43 |
| Frail | No | 2.885 | 2.652 | 3.139 | <0.001 | <0.001 | - | 1.970 | 1.804 | 2.151 | <0.001 | <0.001 | - |
| Frail | Yes | 3.444 | 2.697 | 4.396 | <0.001 | <0.001 | 29.64 | 2.455 | 1.920 | 3.140 | <0.001 | <0.001 | 50.10 |

*Note:* HR = hazard ratio; CI = confidence interval; RD % = percentage risk difference; Ref = reference group; Bonf. = Bonferroni; BH = Benjamini & Hochberg. Age (in years) was used as the underlying time axis. Model 1 – unadjusted; Model 2 – adjusted for sex, ethnicity, highest qualification, Townsend deprivation index, cohabitation with spouse/partner, smoking status, alcohol intake frequency, systolic and diastolic blood pressure, body mass index, cholesterol, multimorbidity count and assessment centre.

**Supplementary Table 11.** All-cause mortality by frailty index categories, no comorbid depression and anxiety disorders

| | | Model 1 | | | | | | Model 2 | | | | | |
|---|---|---|---|---|---|---|---|---|---|---|---|---|---|
| **Depression** | | HR | 95% CI | | $p_{Bonf.}$ | $p_{BH}$ | RD % | HR | 95% CI | | $p_{Bonf.}$ | $p_{BH}$ | RD % |
| Non-frail | No | Ref | - | - | - | - | - | Ref | - | - | - | - | - |
| Non-frail | Yes | 1.082 | 0.969 | 1.208 | >0.999 | 0.180 | - | 1.108 | 0.992 | 1.237 | >0.999 | 0.080 | - |
| Pre-frail | No | 1.372 | 1.319 | 1.427 | <0.001 | <0.001 | - | 1.151 | 1.101 | 1.202 | <0.001 | <0.001 | - |
| Pre-frail | Yes | 1.521 | 1.438 | 1.608 | <0.001 | <0.001 | 39.99 | 1.275 | 1.200 | 1.355 | <0.001 | <0.001 | 82.72 |
| Frail | No | 2.522 | 2.355 | 2.700 | <0.001 | <0.001 | - | 1.598 | 1.476 | 1.730 | <0.001 | <0.001 | - |
| Frail | Yes | 2.849 | 2.613 | 3.106 | <0.001 | <0.001 | 21.47 | 1.736 | 1.576 | 1.912 | <0.001 | <0.001 | 23.03 |
| **Anxiety disorder** | | | | | | | | | | | | | |
| Non-frail | No | Ref | - | - | - | - | - | Ref | - | - | - | - | - |
| Non-frail | Yes | 0.990 | 0.831 | 1.181 | >0.999 | 0.914 | - | 1.036 | 0.869 | 1.235 | >0.999 | 0.731 | - |
| Pre-frail | No | 1.371 | 1.318 | 1.426 | <0.001 | <0.001 | - | 1.151 | 1.101 | 1.203 | <0.001 | <0.001 | - |
| Pre-frail | Yes | 1.286 | 1.179 | 1.402 | <0.001 | <0.001 | -22.97 | 1.143 | 1.045 | 1.250 | 0.069 | 0.004 | -5.03 |
| Frail | No | 2.519 | 2.353 | 2.697 | <0.001 | <0.001 | - | 1.577 | 1.455 | 1.710 | <0.001 | <0.001 | - |
| Frail | Yes | 2.002 | 1.669 | 2.401 | <0.001 | <0.001 | -34.04 | 1.410 | 1.170 | 1.700 | 0.006 | <0.001 | -28.96 |

*Note:* HR = hazard ratio; CI = confidence interval; RD % = percentage risk difference; Ref = reference group; Bonf. = Bonferroni; BH = Benjamini & Hochberg. Age (in years) was used as the underlying time axis. Model 1 – unadjusted; Model 2 – adjusted for sex, ethnicity, highest qualification, Townsend deprivation index, cohabitation with spouse/partner, smoking status, alcohol intake frequency, systolic and diastolic blood pressure, body mass index, cholesterol, multimorbidity count and assessment centre.
